# Endothelial dysfunction determines severe COVID-19 in combination with dysregulated lymphocyte responses and cytokine networks

**DOI:** 10.1101/2021.07.08.21260169

**Authors:** Louisa Ruhl, Isabell Pink, Jenny F. Kühne, Kerstin Beushausen, Jana Keil, Stella Christoph, Andrea Sauer, Lennart Boblitz, Julius Schmidt, Sascha David, Hans-Martin Jäck, Edith Roth, Markus Cornberg, Thomas F. Schulz, Tobias Welte, Marius M. Höper, Christine S. Falk

**Affiliations:** Institute of Transplant Immunology, MHH, D; Department of Pneumology, MHH, D; Department of Intensive Care Medicine, University of Zurich, CH; Division of Molecular Immunology, Internal Medicine III, Nikolaus-Fiebiger-Center of Molecular Medicine, Friedrich-Alexander University Erlangen-Nürnberg, Erlangen, D; Department of Gastroenterology Hepatology and Endocrinology, MHH, D; Center for Individualized Infection Medicine CiiM; Institute of Virology, MHH, D; German Center for Lung Diseases DZL/BREATH; German Center for Infection Research, DZIF, TTU-IICH; Excellence Cluster 2155 RESIST, MHH

**Keywords:** COVID-19, SARS-CoV-2, Endothelium, adaptive immunity, antibodies, cytokines, memory T cells, infection, inflammation, plasma cells

## Abstract

The systemic processes involved in the manifestation of life-threatening COVID-19 and in disease recovery are still incompletely understood, despite investigations focusing on the dysregulation of immune responses after SARS-CoV-2 infection. To define hallmarks of severe COVID-19 and disease recovery in convalescent patients, we combined analyses of immune cells and cytokine/chemokine networks with endothelial activation and injury. ICU patients displayed an altered immune signature with prolonged lymphopenia but expansion of granulocytes and plasmablasts along with activated and terminally differentiated T and NK cells and high levels of SARS-CoV-2-specific antibodies. Core signature of seven plasma proteins revealed a highly inflammatory microenvironment in addition to endothelial injury in severe COVID-19. Changes within this signature were associated with either disease progression or recovery. In summary, our data suggest that besides a strong inflammatory response, severe COVID-19 is driven by endothelial activation and barrier disruption, whereby recovery depends on the regeneration of the endothelial integrity.

## Introduction

In December 2019, patients suffering from severe acute respiratory syndrome (SARS) were observed in Wuhan, China, following infection with the severe acute respiratory syndrome corona virus 2 (SARS-CoV-2), which rapidly spread around the globe and was declared as pandemic by the World Health Organization (WHO) on 11 March 2020. The associated coronavirus disease 2019 (COVID-19) ranges from an asymptomatic state or mild symptoms to severe progression and lethal outcome. As of May 2021 SARS-CoV-2 infected more than 150,000,000 people worldwide and caused more than 3,200,000 deaths.

The upper respiratory tract and the lung are the primary sites of SARS-CoV-2 infection and dyspnea is a leading symptom of severe COVID-19 with respiratory failure causing admission to intensive care unit (ICU) and mechanical ventilation (1). Pulmonary symptoms are mainly caused by infection of epithelial cells, particularly type II alveolar epithelial cells, via Angiotensin-converting enzyme 2 (ACE2) as receptor for the SARS-CoV-2 spike protein following proteolytic cleavage by the TMPRSS2 protease, facilitating viral entry (2). Sustained uncontrolled infection results in extensive death of barrier cells within the alveoli and leads to vascular leakage along with tissue edema and activation of coagulation pathways (3). These processes ultimately promote the formation of an acute respiratory distress syndrome (ARDS), a major clinical feature of severe COVID-19. However, in severe cases, development of heart diseases, abnormal blood coagulation and neurological complications as well as liver and kidney injuries are also associated with COVID-19 and can progress to multi-organ failure (4). These clinical symptoms indicate a crucial participation of the endothelial system in the manifestation of severe COVID-19.

The reasons for the wide range of COVID-19 symptoms and variable, unpredictable disease progressions are under intense investigation. Age and age-related chronic inflammatory conditions such as diabetes together with genetic predispositions represent major risk factors for severe COVID-19 (5). Furthermore, several studies suggested a direct connection between the host immune response and critical COVID-19, demonstrating dynamic and highly heterogeneous immune signatures with altered immune cell compositions and cytokine/chemokine patterns in severe COVID-19 (6-9). SARS-CoV-2 seems to manipulate the host immunity by evading especially the innate immune response, as ineffective IFN immunity was associated with fatal COVID-19 (5,10). Besides the innate, also the adaptive immunity contributes to progression towards severe COVID-19 (10). In particular, SARS-CoV-2 specific T cells are crucial for rapid virus clearance, since an extended absence of virus-specific T cells was related to severe COVID-19 progression (8). Regarding the humoral response, >90% of infected individuals seroconverted to SARS-CoV-2 spike (S) and/or nucleocapsid (N) proteins, but rather limited somatic hypermutation was observed for neutralizing antibodies (7,11), indicating also an altered B cell response. Additionally, cytokine profiles from severe SARS-CoV-2 infections revealed a highly inflammatory microenvironment with elevated levels of cytokines like IL-6 and TNF-*α* and chemokines like CXCL-10 in addition to altered antiviral IFN-responses (5,6,12).

As clinical symptoms indicate a crucial endothelial contribution to severe COVID-19 our study aimed to investigate a potential link between the endothelium and hyperinflammation in critical cases. We focused on identifying immune and endothelial signatures as main drivers for severe COVID-19 progression since uncovering pathological mechanisms may be a first step for developing novel treatment strategies and could help to understand long-lasting effects in recovered patients (13). Therefore, we included blood samples from acute severe COVID-19 patients admitted to intensive care unit (ICU, n=58) and from convalescent, former ICU patients (CONV, n=28) in our study. Analyzing cellular, humoral and endothelial responses by unsupervised clustering and correlation analyses of 98 immune cell subsets, 83 soluble plasma proteins and S-and N-specific IgM, IgA and IgG antibodies, we observed a dominating hyperactivated HLA-DR^+^/CD38^+^ phenotype of memory T and NK cells despite massive T cell lymphopenia. Furthermore, granulocytes were significantly expanded as well as plasmablasts which were accompanied by a rapid production of S-and N-specific IgM, IgA and IgG antibodies. Third, in severe COVID-19, this hyperactivation at cellular level was accompanied by a highly pro-inflammatory cytokine/chemokine network, which was associated with endothelial activation leading to a stepwise disruption of the endothelial barrier. Finally, unsupervised cluster and correlation analyses identified a unique core chemokine/endothelial signature for severe COVID-19 as well as distinct cytokine signatures associated with recovery from severe disease.

## Results

### Dynamic changes in the entire immune cell composition in severe COVID-19 patients with memory T cell development and plasmablast expansion

A total of N=42 patients with n=86 samples from MHH with confirmed SARS-CoV-2 infection were analyzed at various time points after symptom onset. N=25 patients (n=58 samples) have been admitted to ICU and N=17 patients (n=28 samples) were convalescent former ICU patients (CONV). N=29 unexposed individuals (UE, n=36) served as control group (Suppl. Tab. 1). To analyze the leukocyte dynamics during COVID-19, we quantified their absolute numbers and proportions in ICU, CONV and UE. Unsupervised cluster analyses of our immune phenotyping data sets revealed a clear separation of ICU from CONV and UE, indicating significant dynamic changes in the lymphocyte compartment during acute COVID-19 (Fig. 1 A-B, S1A). As known from other studies (14,15), both absolute numbers as well as frequencies of granulocytes were highly elevated in blood of ICU patients while monocytes were stable and only proportionally reduced (Fig. 1C, S1B). In contrast, absolute numbers as well as frequencies of lymphocytes, especially T cells, were significantly decreased in ICU compared to CONV and UE samples, with some patients showing strong lymphopenia (<1000 lymphocytes/µl blood). The number of NK cells was also decreased, whereas B cell numbers were unchanged in ICU compared to CONV and UE. Leukocyte numbers and frequencies of CONV displayed no significant differences compared to UE (Fig. C, S1B). Interestingly, four out of five deceased ICU patients clustered close to each other and exhibited decreased numbers of T cells and an activated immune phenotype (Fig. 1B, S1A). As reported earlier, ICU patients show a strong decline in lymphocyte numbers, including T and NK cells, leading to lymphopenia along with an unusual expansion of granulocytes. Furthermore, comparable leukocyte numbers and frequencies of CONV and UE indicate recovery at the cellular level in the periphery of CONV.

**Fig. 1:**
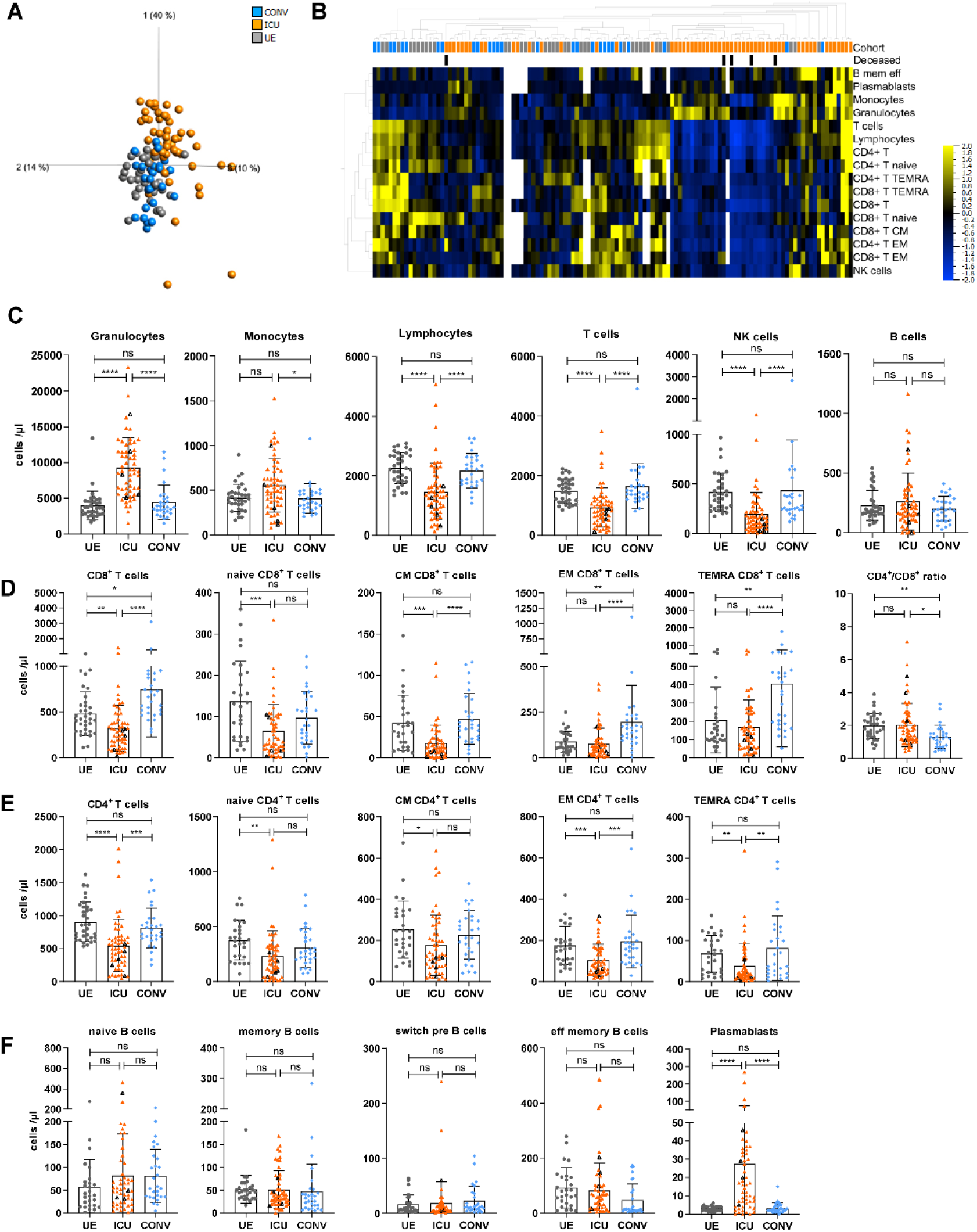
Dynamic changes in the entire immune cell composition in severe COVID-19 patients with memory T cell development and plasmablast expansion. Immune cell distribution reflected by absolute numbers in patient blood was analyzed using TruCount analyses. **(A)** Principle component analysis. Multigroup comparison with a p-value cut off of 0.041 was used to identify significant differences among UE (n=36), ICU (n=58) and CONV (n=28). **(B)** Heatmap analysis. Multigroup comparison with a p-value cut off of 0.041 was used to identify significant differences among UE (n=36), ICU (n=58) and CONV (n=28). **(D-F)** Numbers of different immune cells in patient blood. T cells: naïve (CCR7+CD45RO-), central memory (CM, CCR7+CD45RO+), effector memory (EM, CCR7-CD45RO+) and TEMRA (CCR7-CD45RO-); B cells: naïve (IgD+CD27-) memory (mem, CD27+IgD-), switch precursor (switch pre, CD27+IgD+), effector memory (eff mem, IgD-CD27-) and plasmablasts (CD19+CD20-CD27+CD38+) Black triangles represent last samples from deceased patients. UE: Unexposed donors, ICU: Intensive care unit patients, CONV: convalescent patients, CM: central memory, EM: effector memory, mem eff: memory effector Statistical analysis: ANOVA test with Turkey multiple comparison test or Kruskal-Wallis with test with Dunn’s multiple comparison test were performed. *p < 0.05, **p<0.01, ***p<0.001, ****p<0.0001.

To further investigate the T cell phenotypes in severe COVID-19, the distribution of naïve vs. memory CD4^+^ and CD8^+^ T cells was determined for ICU, CONV and UE based on expression of the chemokine receptor CCR7 and the CD45RO memory marker (Fig. S2A). Especially ICU showed highly variable numbers and proportions of all T cell subsets. CD8^+^ T cell numbers, particularly CCR7^+^CD45RO^-^ naïve and CCR7^+^CD45RO^+^ central memory (CM) T cells were decreased in ICU compared to CONV and UE (Fig. 1D). Also, all CD4^+^ T cells subsets were significantly reduced in numbers (Fig. 1E). With regard to CONV, we found elevated numbers of CD8^+^ T cells compared to UE, especially CCR7^-^CD45RO^+^ effector memory (EM) and CCR7^-^CD45RO^-^ TEMRA at the cost of naïve CD8^+^ T cells, which translated into a reduced CD4^+^/CD8^+^ ratio compared to UE and ICU (Fig. 1D). Thus, CD4^+^ and CD8^+^ T cells were both affected by the lymphopenia in COVID-19 ICU patients and particularly elevated CD8^+^ TEMRA T cell numbers in CONV suggest a pronounced T cell differentiation during SARS-CoV-2 infection.

In contrast to T cells, B cells were less affected by the lymphopenia in ICU (Fig. 1C, F). Detailed analyses of B cell subsets showed a broad range of IgD^+^CD27^-^ naïve, CD27^+^IgD^-^ memory (mem), CD27^+^IgD^+^ switch precursor (switch-pre) and IgD^-^CD27^-^ effector memory (eff-mem) B cells in ICU accompanied by an unusual expansion of CD19^+^CD20^-^CD27^+^CD38^+^ plasmablasts (Fig. 1F, S1E, S2C). Even though we did not observe significant differences in absolute numbers of these B cell subsets in ICU, frequencies of naïve B cells were increased and frequencies of eff-mem B cells were decreased in CONV compared to UE and ICU (Fig. 1F, S1E). Concluding, elevated absolute numbers and proportions of plasmablasts in ICU patients indicate a strong systemic B cell response during acute SARS-CoV-2 infection followed by a reverse transformation of the B cell population in CONV with elevated proportions of naïve B cells and reduced proportions of eff-mem B cells.

### Plasmablasts contribute to antibody development and blocking activity is associated with disease severity

Based on the massive plasmablast expansion in ICU, we hypothesized that their presence in circulation might be accompanied by the appearance of SARS-CoV-2-specific antibodies. A Luminex-based multiplex assay was used for the detection of IgM, IgA and IgG specific for the S1, the receptor binding domain (RBD), the S2 regions of the spike protein (S) and the nucleocapsid (N) antigen of SARS-CoV-2 in plasma of ICU and CONV, while UE served as controls. In ICU, high IgG relative concentrations (MFI) were detected against all four SARS-CoV-2 antigens with the exception of two B cell-depleted patients, while IgM and IgA displayed broad variations of 45-100% positive samples (Fig. 2A-C). In contrast, CONV showed decreased proportions of seroconversion and lower concentrations of IgM and IgA (Fig. 2A-D). Since the ICU samples were obtained during acute infection, they were collected earlier in the course of disease than CONV samples, which was reflected by the different IgM, IgA and IgG kinetics and thus illustrated a time-dependent antibody class-switch from IgM to IgG and IgA (Fig. S3A, B). The decline of IgM and IgA levels 20 days after symptom onset (DASO) in ICU was accompanied by increased IgG levels that remained high up to 200 days after symptom onset in CONV (Fig. S3A, B). No major differences in IgM, IgA and IgG responses were detected between the S-and N-antigens arguing for humoral responses against multiple SARS-CoV-2 antigens (Fig. 2D).

**Fig. 2:**
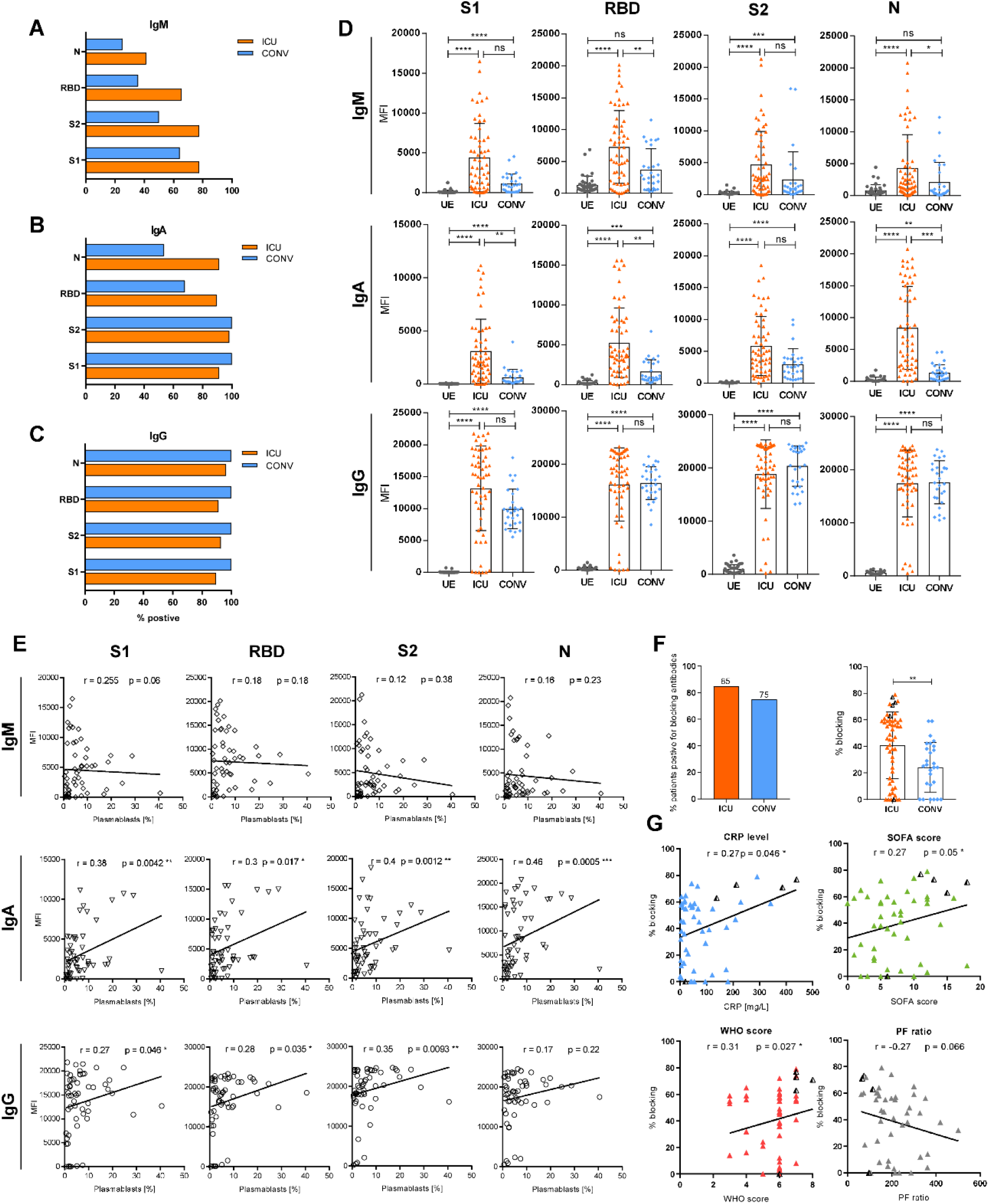
Plasmablasts contribute to antibody development and blocking activity is associated with disease severity. **(A-E)** Luminex-based multiplex assay was used to detect IgM, IgA and IgG antibodies against S1-, RBD, S2-, or N-antigen of SARS-CoV-2 in patient sera. **(A-C)** Percentage of seroconverted ICU (n=58) and CONV (n=28) patients for SARS-CoV-2-specific IgM (A), IgA (B) and IgG (C) antibodies. Threshold for positive samples was calculated based on the mean fluorescent intensity (MFI) of antibodies from 36 UEs + 2x standard deviation. **(D)** Antibody levels from UE (n=36), ICU (n=58) and CONV (n=28) are displayed as MFI. **(E)** Correlation analysis between S-and N-specific antibodies and plasmablasts proportions. **(F)** Percentage of ICU (n=58) and CONV (n=28) which developed blocking antibodies against SARS-CoV-2 RBD (left) and efficacy of blocking activity (right), which was assessed by competitive ELISA. Efficient blocking was expressed as the percent blocking at a 1:50 serum dilution relative to a UE serum control. **(G)** Correlation analysis between COVID-19 severity markers (CRP levels, SOFA-, WHO-score, PF ratio) and blocking efficiency. Black triangles represent last samples from deceased patients. Statistical analysis: Multigroup comparisons were performed using ANOVA test with Turkey multiple comparison test or Kruskal-Wallis with test with Dunn’s multiple comparison test. Two group comparison was performed using Mann-Whitney test; Spearman-Correlation. *p < 0.05, **p<0.01, ***p<0.001, ****p<0.0001.

To unravel a potential relationship between plasmablast expansion and the occurrence of S-or N-specific antibodies, we correlated the proportions of plasmablasts with the corresponding relative antibody levels. While no correlation was found between plasmablasts and IgM antibodies, there was a weak positive correlation between plasmablasts and S-and N-specific IgA and IgG (Fig. 2E). Moreover, a competitive ELISA was performed to analyze whether antibodies in ICU were able to block *in vitro* binding of SARS-CoV-2 RBD to the ACE2 receptor. Interference was calculated as percent blocking in the presence of ICU plasma (1:50 dilution) relative to UE control plasma without SARS-CoV-2 antibodies. Interestingly, 85% of ICU and 75% CONV plasma samples contained interfering antibodies (Fig. 2F), but the blocking efficacy of ICU antibodies was significantly higher (median 40%) than CONV antibodies (median 24%) (Fig. 2F). Especially deceased patients exhibited high blocking activity (>60%) except for one B cell-depleted patient. Moreover, we correlated antibody blocking efficacy with disease severity makers: C-reactive protein (CRP) level as a marker for systemic inflammation, sepsis-related organ failure assessment (SOFA) score for the extent of systemic organ failure, World Health Organizations (WHO) eight-point scale for COVID-19 trial endpoints (https://www.who.int/publications/i/item/covid-19-therapeutic-trial-synopsis) and PaO2/FiO2 (PF) ratio representing pulmonary function. CRP levels, SOFA-and WHO-scores showed weak positive correlations with the antibody-blocking efficacy in ICU, whereas PF ratios were negatively correlated. Interestingly, deceased patients displayed remarkably high CRP-levels, SOFA-and WHO-scores and low PF ratios (Fig. 2G).

These results illustrate that the majority of ICU developed rapid and sustained IgG antibody responses specific for SARS-CoV-2 S-and N-proteins and their blocking efficacy seemed to increase with disease severity suggesting that antibodies were insufficient to protect from developing severe COVID-19.

### Expansion of activated and terminally differentiated T and NK cells in COVID-19 ICU patients

Next, we aimed to identify the major immune cell populations significantly altered in ICU versus UE to determine ICU-associated cellular patterns. Unbiased volcano plot analysis depicted a strong decline in lymphocytes but high frequencies of HLA-DR^+^CD38^+^ activated CD4^+^ and CD8^+^ T cells together with HLA-DR^+^ NK cells and CD57^+^ terminally differentiated T and NK cells in ICU compared to UE (Fig. 3A, B). These high frequencies of activated HLA-DR^+^CD38^+^ T cells (Fig. 3C) together with HLA-DR^+^ and CD69^+^ activated CD56^dim^ NK cells (Fig. 3D) were associated with acute COVID-19 and decreased with recovery in CONV (Fig. 3E, F). Proportions of CD28^-^CD27^-^ or CD57^+^CCR7^-^ terminally differentiated T and CD57^+^ NK cells were increased in ICU and remained elevated in CONV compared to UE (Fig. 3F). On the other hand, CD127^+^CD8^+^ T cells frequencies were decreased in ICU as well as in CONV compared to UE. Thus, severe SARS-CoV-2 infection is associated with strong activation and differentiation into late memory T and NK cells.

**Fig. 3:**
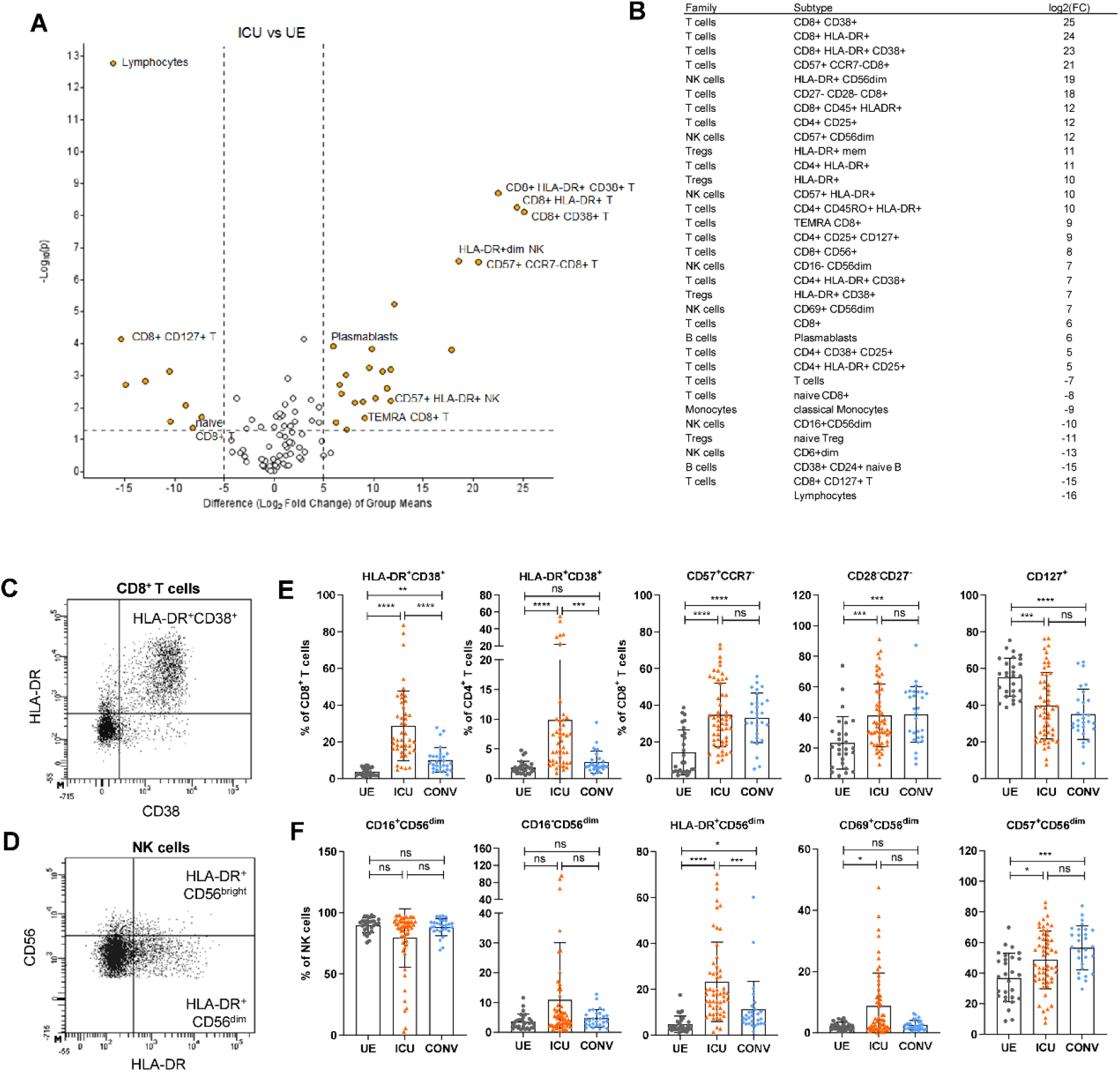
Expansion of activated and terminally differentiated T and NK cells in COVID-19 ICU patients. (**A**) Volcano Plot visualizing two-group comparison of flow cytometry data including proportions of immune cells from ICU (n=58) and UE (n=36). (**B**) Significantly (p ≥ 0.05) altered immune cell subsets between ICU (n=58) and UE (n=36) ordered based on their fold-change. **(C, D)** Representative flow cytometry plots **(E, F)** Representative flow cytometry data of immune cell frequencies from UE (n=36), ICU (n=58) and CONV (n=28). Statistical analysis: Multigroup comparison was performed using ANOVA test with Turkey multiple comparison test or Kruskal-Wallis with test with Dunn’s multiple comparison test; *p < 0.05, **p<0.01, ***p<0.001, ****p<0.0001.

### In severe COVID-19 lymphocyte dynamics correlate with disease progression

In order to examine the implications of the significant changes within the lymphocyte compartment in ICU, they were correlated to disease severity (SOFA-, WHO-score, CRP level) and progression (days after symptom onset DASO). Several immune cell subsets significantly correlated with DASO, for example plasmablasts with a negative correlation, showing that plasmablasts developed early during severe disease but decreased with prolonged ICU hospitalization (Fig. 4A, B). In the myeloid compartment, proportions of CD14^+^CD16^+^ intermediate and CD14^-^CD16^+^ non-classical monocytes showed a positive correlation to DASO. In parallel, NK cells increased with a simultaneous reduction of activated CD69^+^ or CD25^+^ NK cells over time (Fig. 4C, D). Moreover, different T cell subsets correlated with disease duration, indicating a time-dependent T cell differentiation caused by severe COVID-19 (Fig. 4E). While naïve CD8^+^, CD4^+^ T and CD25^hi^CD127^lo^ Treg cells decreased after symptom onset, EM as well as HLA-DR^+^CD38^+^ activated CD8^+^ T cells accumulated over time (Fig. 4E, F). As expected, frequencies of naïve CD8^+^ T cells negatively and EM CD8^+^ T cells positively correlated with patient age in ICU (Fig. S4A, B). However, since no correlation was observed between COVID-19 disease duration and patient age we conclude that patient age had a minor impact on T cell differentiation during severe COVID-19 (Fig. S4C). Remarkably, CD4^+^ T cells and CD8^+^ TEMRA T cells did not follow the usual age correlation in ICU compared to UE (Fig. S4 A, B).

**Fig. 4:**
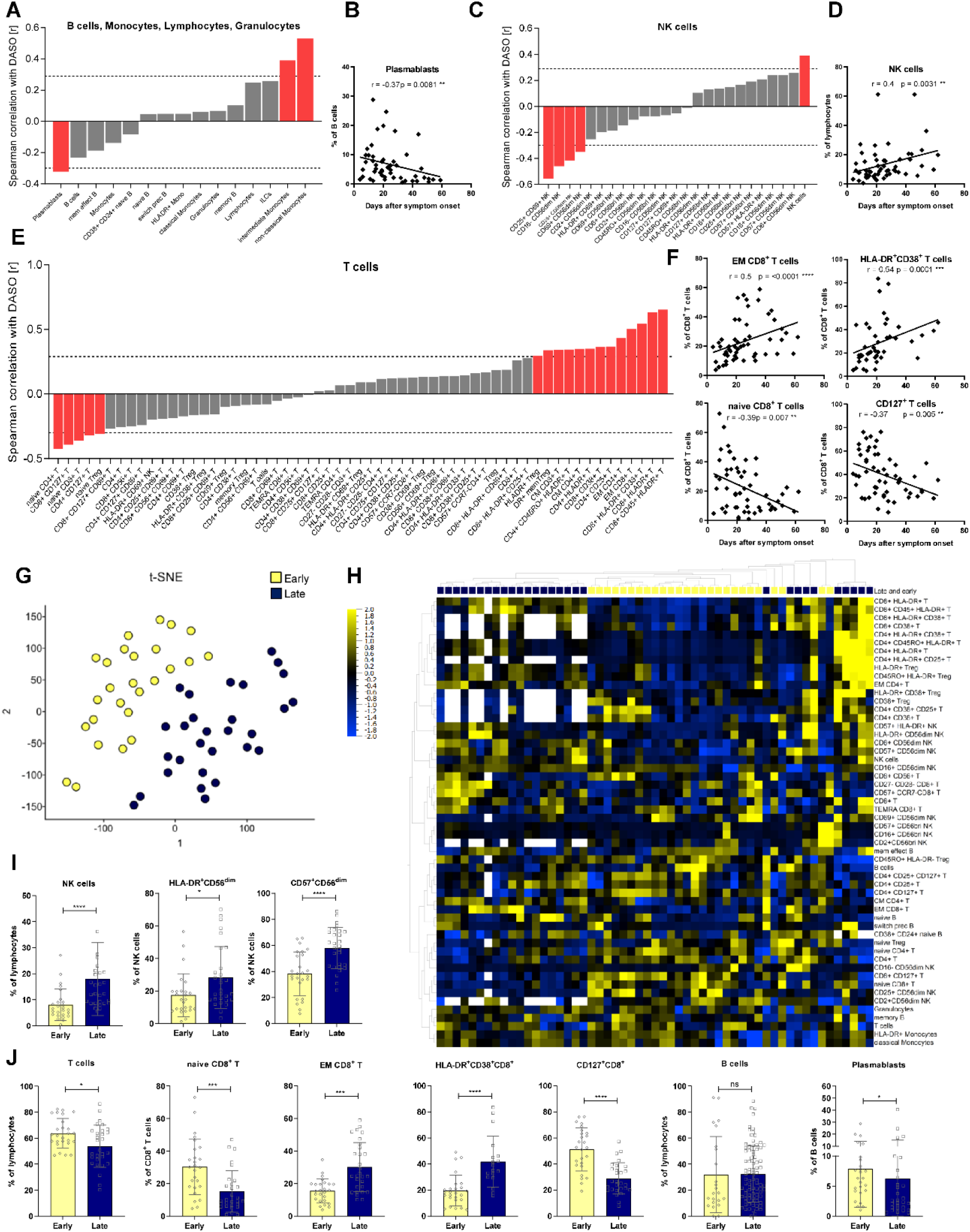
In severe COVID-19, lymphocyte dynamics correlate with disease progression. **(A, C, E)** Waterfall-plot representing correlation coefficient (r) of Spearman-correlation analysis between immune cell proportions of B cells, monocytes, lymphocytes and granulocytes (A) or NK cells (C) or T cells (E) and disease duration as days after symptom onset (DASO) from ICUs (n=58). Red columns represent significant results. Dotted lines represent the minimal Spearman correlation coefficient (r) required for significant correlation **(B, D, F)** Spearman correlation analysis between proportions of plasmablasts (B), NK cells (D) or different T cell populations (F) and DASO from ICUs (n=58). **(G)** tSNE analysis of flow cytometry data from ICU (n=58). Variance-value cut off of 0.305 was used to identify significant differences within ICU cohort. Patients were classified into two groups. Blue: Late subgroup (n=32), yellow: early subgroup (n=26). **(H)** Heatmap of flow cytometry data including 98 immune cell populations from ICU (n=58). Variance-value cut off of 0.035 was used to identify significant differences within ICU cohort. Samples and immune cell subsets were ordered according to hierarchical clustering. Blue to yellow scale indicates the prevalence of each subset. Missing values are displayed in white. **(I-J)** Representative immune cell proportions from early (n=26) and late patients (n=32) within ICU cohort. Statistical analysis: Unpaired t-test or Mann-Whitney test; Spearman-Correlation. *p < 0.05, **p<0.01, ***p<0.001, ****p<0.0001.

Unlike disease progression, severity markers (SOFA-, WHO-score, CRP level) correlated with only a few immune cell subsets, suggesting that disease severity had only a minor effect on the immune cell distribution (Fig. S5A). As expected, especially activated immune cells like CD69^+^CD8^+^ T cells and CD69^+^CD56^bright^ NK cells positively correlated with CRP levels.

Furthermore, t-distributed stochastic neighbor embedding (tSNE) (Fig. 4G) and unsupervised cluster analyses (Fig. 4H) revealed two distinct subgroups of ICU samples, which separated based on their DASO into a “late” and “early” subgroup (Fig. 4H, S5B, S5C). The “late” ICU subgroup consisted of samples obtained between day 7 to 68 (mean 34 days), whereas the “early” ICU subgroup consisted of samples from day 4 to 29 after symptom onset (mean 15 days). Neither of the two groups displayed significant differences regarding their SOFA-and WHO-scores nor PF ratios but the late subgroup exhibited decreased CRP levels compared to the early subgroup (Fig. S5D). However, detailed analyses of the immune cell compositions illustrated that these two subgroups could additionally be distinguished by several immune cell proportions (Fig. 4H-J). Particularly expression of the activation markers HLA-DR, CD38 and CD25 and frequencies of CD45RO^+^ or CD57^+^ memory T and NK cells increased with disease duration in ICU, whereas consequently, proportions of naïve T and NK cells declined over time (Fig. 4H-J).

These results show that the alterations within the lymphocyte compartment are strongly time-dependent during the individual ICU period but are less directly associated with disease severity.

### Systemic inflammation and endothelial injury in COVID-19 patients

With regards to the systemic microenvironment, we quantified concentrations of 83 plasma proteins in ICU, CONV and UE including cytokines, chemokines, growth factors and endothelial factors. Samples from ICU clearly clustered separately from CONV and UE samples and displayed significantly increased levels of 65 plasma proteins (Fig. 5A). Deceased ICU patients clustered in separated patterns, indicating different plasma protein signatures associated with fatal outcome. To identify the most statistically significant plasma proteins discriminating ICU from UE, we analyzed their levels based on the fold change between ICU and UE (Fig. 5B). The volcano plot illustrates a COVID-19-associated plasma protein profile composed of pro-inflammatory cytokines and chemokines (e.g. IL-6, CXCL10) in addition to growth factors (e.g. granulocyte-macrophage colony-stimulating factor (GM-CSF)) and endothelial factors (e.g. osteopontin (OPN), angiopoietin-2 (Ang-2)). In particular, chemokines and endothelial activation markers correlated with each other in clusters (Fig. 5C), indicating a co-regulated secretion and suggesting that inflammation and endothelial injury may collectively contribute to severe COVID-19. Furthermore, we observed a high variability in the respective plasma proteins levels within the ICU cohort, as previously observed for immune cell populations (Fig. 5D, S6). In addition to classical inflammation-associated cytokines (e.g. IL-1*β*, IL-6, TNF-*α*, IFN-*γ*) ICU displayed increased levels of soluble interleukin-6 receptor α (sIL-6Rα), urokinase-type-plasminogen-activator (uPA) and its antagonist plasminogen activator inhibitor 1 (PAI-1) (Fig. 5D, S6). Whereas Ang-2, a central mediator of endothelial activation, was elevated, the concentrations of its soluble angiopoietin receptor 2 (sTIE-2) were stable in ICU, CONV and UE, suggesting a regulation at the ligand and not the receptor level. Endothelial activation in ICU was further detectable by significantly higher IGFBP-1, OPN and VCAM levels, accompanied by several others. Organ contribution could be seen by higher hepatocyte growth factor (HGF) levels, for instance (Fig. 5D). Of note, the plasma protein profile of CONV displayed no significant alterations compared to UE.

**Fig. 5:**
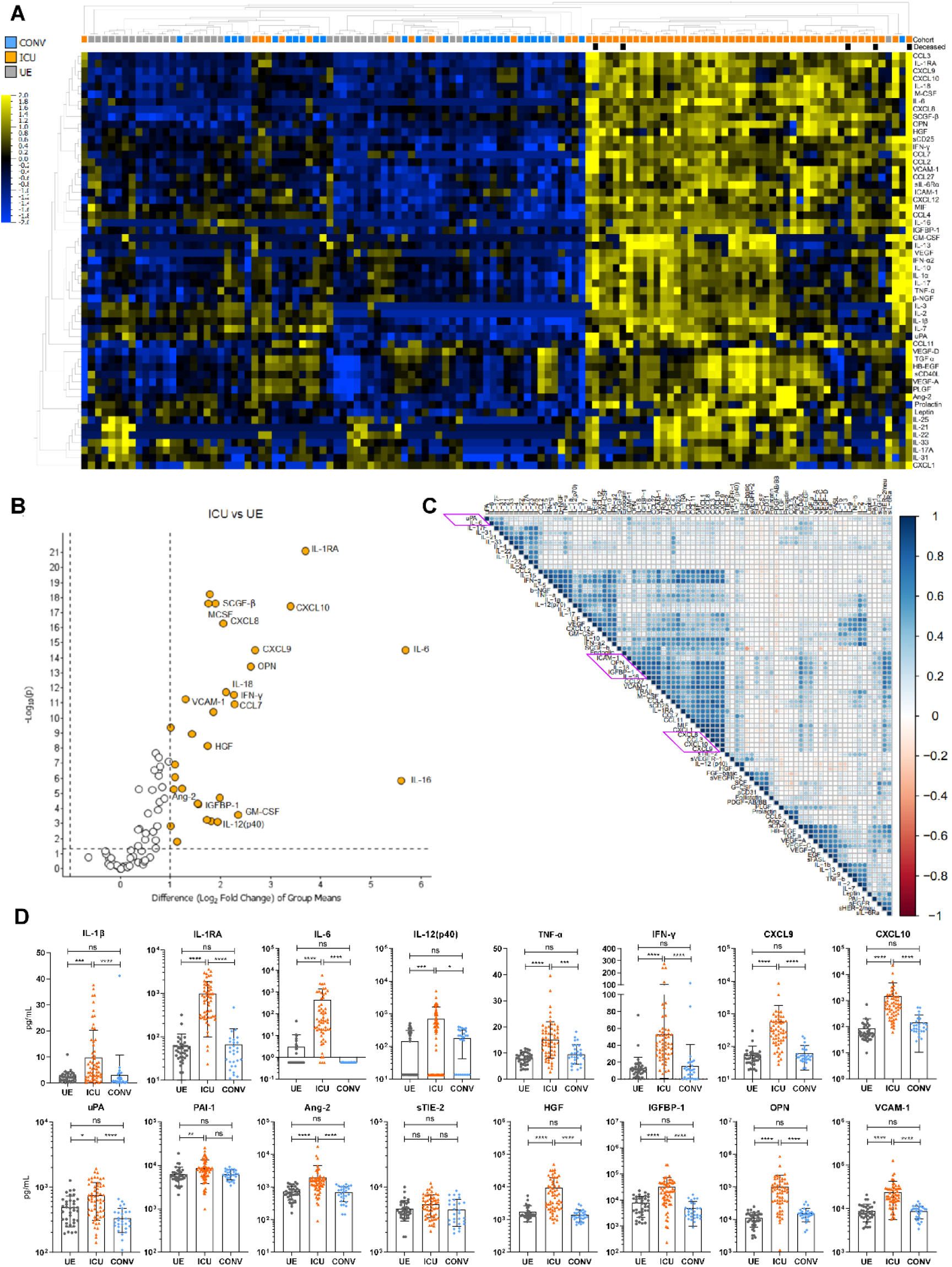
Systemic inflammation and endothelial injury in COVID-19 patients. Cytokine concentrations in patient sera were measured by Luminex-based multiplex assay **(A)** Heatmap of cytokine data including 83 plasma proteins from UE (n=36), ICU (n=58) and CONV (n=28). P-value cut off of 0.01 was used to identify significant differences among the three cohorts. Samples and immune cell subsets were ordered using to hierarchical clustering. Blue to yellow scale indicates the prevalence of each subset. Missing values are displayed in white. **(B)** Volcano Plot visualizing a two-group comparison of plasma protein data from ICU (n=58) and UE (n=36). (**C**)Correlation matrix of 83 plasma proteins from ICUs. Spearman correlation was used to calculate correlation coefficients, which were displayed in circles. Strength of correlation was depicted by circle size. Cytokines were ordered using hierarchical clustering. Red to blue scale indicates the prevalence of each subset. **(D)** Representative cytokine concentrations from UE (n=36), ICU (n=58) and CONV (n=28). Statistical analysis: Kruskal-Wallis with test with Dunn’s multiple comparison test were performed. *p < 0.05, **p<0.01, ***p<0.001, ****p<0.0001.

In summary, we observed a highly inflammatory profile of plasma proteins in ICU, indicating strong immune system activation due to SARS-CoV-2 infection. Furthermore, high levels of endothelial factors suggest that endothelial injury may be a crucial contributor to severe COVID-19 progression.

### Core plasma protein signature of severe COVID-19 consists of eleven inflammatory mediators and endothelial factors

Based on the extensive secretion of immunomodulating proteins and endothelial factors in ICU, we aimed to examine their implications for disease severity (SOFA-, WHO-scores, CRP levels) and disease duration (DASO). Several plasma protein levels positively correlated with disease duration such as IL-13, IL-1*β*, GM-CSF and the vascular endothelial growth factor (VEGF) showing an extraordinary increase beyond day 20 after symptom onset (Fig. 6A, B). In contrast, CXCL10 and IL-12(p40) concentrations significantly decreased over time. Deceased ICU patients displayed particularly low IL-1β and IL-13 levels, while CXCL10 and IL-12(p40) plasma concentrations were increased (Fig. 6B). High SOFA scores correlated with high levels of chemokines like CXCL10 and endothelial factors like HGF, insulin-like growth factor-binding protein 1 (IGFBP-1) and uPA (Fig. 6C, D). Based on the highly inflammatory profiles of these ICU patients, it was not surprising that correlation analyses with CRP levels and WHO scores showed almost identical patterns (data not shown). Comparing plasma proteins positively correlating to either CRP levels, SOFA-or WHO-scores, we detected an intersection of seven shared plasma proteins, representing the core signature associated with severe COVID-19 (Fig. 6E). This signature was composed of the pro-inflammatory cytokines IL-12(p40) and IL-6, the growth factor stem cell growth factor beta (SCGF-*β*), the chemokines CXCL8-10 and the endothelial factor HGF. Furthermore IGFBP-1, IL-16, M-CSF and VCAM-1 positively correlated with SOFA-and WHO-scores, representing the contribution of the endothelium to severe COVID-19. Additional plasma proteins as IL-17F, IL-25 and CCL2 contributed to the unique WHO pattern, while IL-31, CCL27, CCL7, uPA and sCD25 were included to the unique SOFA score-associated pattern and CXCL1 completed the CRP pattern.

**Fig. 6:**
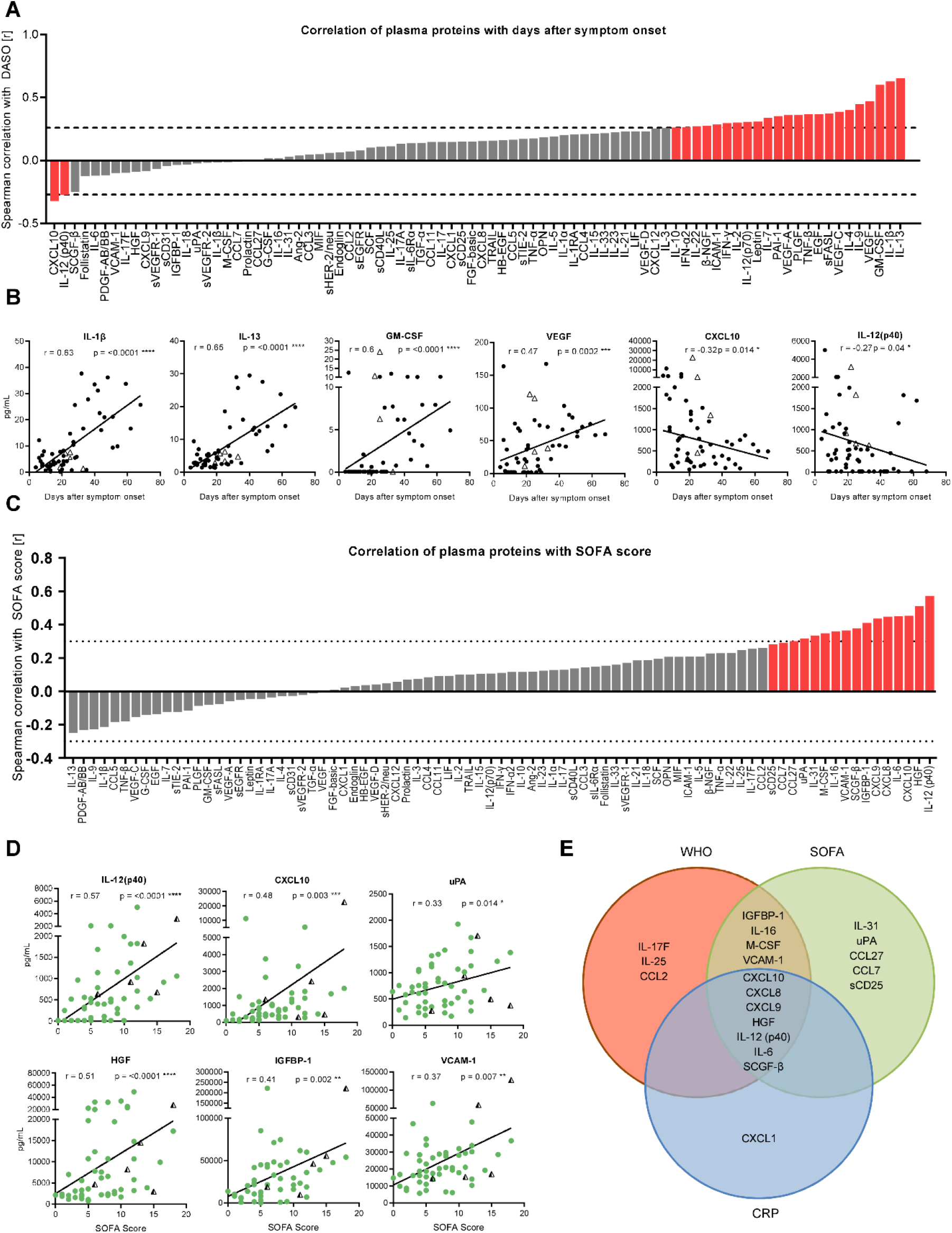
Core plasma protein signature of severe COVID-19 consists of eleven inflammatory mediators and endothelial factors. Cytokine concentrations in patient sera were measured by Luminex-based multiplex assay **(A, C)** Waterfall-plot representing correlation coefficient (r) of Spearman-correlation analysis between cytokines and disease duration as DASO **(A)** or SOFA score **(C)** from ICUs (n=58). Red columns represent significant results. Dotted lines represent the minimal Spearman correlation coefficient (r) required for significant correlation. **(B)** Spearman correlation analysis between representative cytokines and DASO. **(D)** Spearman correlation analysis of representative cytokines with SOFA score. **(E)** Venn-diagram displaying significantly positive correlations between plasma proteins and CRP levels, SOFA-and WHO-score. Triangles represent last samples from deceased patients. Statistical analysis: Spearman-Correlation. *p < 0.05, **p<0.01, ***p<0.001, ****p<0.0001.

Taken together, these signatures demonstrate that severe COVID-19 is characterized by extensive secretion of multifunctional interleukins, growth factors and endothelial factors into the circulation, suggesting that SARS-CoV-2 causes more than a systemic inflammation but multiple endothelial injuries.

### Endothelial dysregulation as major contributor to severe COVID-19

Finally, since we observed distinct plasma protein patterns in deceased ICU patients (Fig. 5A), we wanted to assess whether severe COVID-19 progression could be associated with distinct profiles of ICU patients. Unsupervised t-SNE analysis of plasma proteins revealed three different subgroups within the ICU cohort (Fig. 7A) that clearly clustered separately, with the highest cytokine concentrations in subgroup 3 (Sub3) followed by subgroup 2 (Sub2) and subgroup 1 (Sub1) (Fig. 7B). Furthermore, Sub1-3 differed by their disease severity (SOFA-, WHO-scores, CRP levels, PF ratio) and disease progression (DASO) (Fig. 7C, D). Of note, Sub2 contained ICU patients (n=16) with the lowest CRP levels, SOFA-and WHO-scores but highest PF ratios and with the longest disease durations (DASO) plus one deceased patient with chronic renal failure after renal transplantation (Fig. 7C, D). On the contrary, Sub3 (n=28) displayed the highest SOFA-and WHO-scores among the three subgroups containing two deceased patients. Lowest PF ratios and highest CRP-levels were observed for Sub1 (n=14) with two deceased patients. The plasma protein profiles demonstrated clear differences among these three ICU subgroups (Fig. 7E). In general, lowest cytokine levels were observed for Sub1, which nevertheless still displayed significantly higher levels of key cytokines and chemokines like IL-6, IFN-*γ* and CXCL10 compared to UE (Fig. S7). Sub3 exhibited the strongest inflammatory signature with high levels of pro-inflammatory cytokines like IL-6, IL-18 and TNF-*α* in addition to sIL-6R*α* and the chemokines CXCL8-10 (Fig. 7D, S7). Remarkably, Sub3 had the highest levels of uPA, Ang-2, HGF, IGFBP-1, OPN, ICAM-1 and VCAM-1 demonstrating endothelial activation, damage and multi-organ failure (Fig. 7D, S7). Most characteristic for Sub2 was an extensive secretion of IL-1*β*, IL-13, GM-CSF and VEGF (Fig. 7D, S7), which was already observed as “late” ICU signature (Fig. 6B). Compared to Sub3, Sub2 exhibited significantly lower IL-6 and IL-16 levels but concentrations of IL-18, TNF-*α*, Ang-2 and OPN were still elevated compared to Sub1 and UE (Fig. 7D, S7). However, concentrations of tissue factors like IGFBP-1, uPA and HGF were comparable to UE, suggesting less endothelial dysfunction in Sub2 (Fig. S7).

**Fig. 7:**
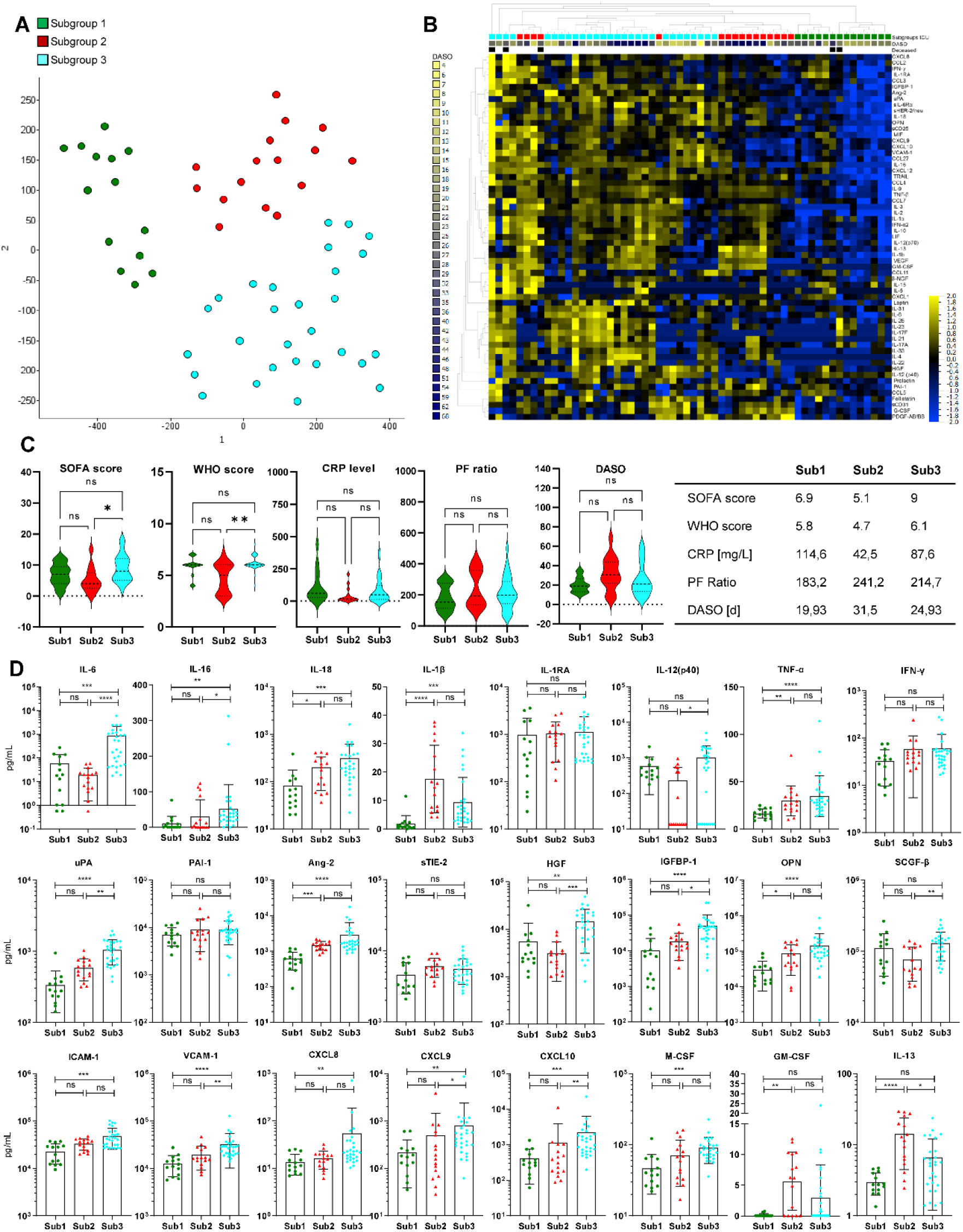
Endothelial dysregulation as major contributor to severe COVID-19. Plasma protein concentrations in patient sera were measured by Luminex-based multiplex assay **(A)** tSNE analysis of 83 plasma protein data from ICU (n=58). Variance-value cut off of 0.146 was used to identify significant differences within ICU cohort. Patients were classified into three subgroups. Green: subgroup 1 (sub1), red: subgroup 2 (Sub2), blue: subgroup 3 (Sub3). **(B)** Heatmap of 83 plasma protein from ICU (n=58). Variance cut off of 0.146 was used to identify significant differences among the three cohorts. Samples and cytokines were ordered using hierarchical clustering. Blue to yellow scale indicates the prevalence of each subset. Missing values are displayed in white. **(C)** Distribution of severity markers SOFA, WHO-score, CRP levels, PF ratio and disease duration (DASO) among the three ICU subgroups. **(D)** Mean-values of SOFA-, WHO-score, CRP level, PF ratio and DASO for the three ICU subgroups. **(E)** Representative cytokine concentrations from ICU subgroups Sub1 (n=14), Sub2 (n=16) and Sub3 (n=28). Statistical analysis: ANOVA test with Turkey multiple comparison test or Kruskal-Wallis with test with Dunn’s multiple comparison test were performed. *p < 0.05, **p<0.01, ***p<0.001, ****p<0.0001.

In summary, the plasma protein profile of Sub3 implies massive inflammation, endothelial injury and impairment of multiple organs, which is further supported by their high SOFA-and WHO-scores. Patients from Sub2 had longer disease durations and decreased SOFA-and WHO-scores compared to patients from Sub1 and Sub3. Together with decreased pro-inflammatory markers and endothelial factors, this indicates a gradual recovery from COVID-19 for Sub2 patients. Lastly, based on their plasma protein profile and clinical data, Sub1 patients are considered as patients at the beginning of a severe COVID-19 progression with massive inflammation but restricted endothelial damage and limited impairment of multiple organs.

## Discussion

Despite the intense global COVID-19 research, it is still incompletely understood which molecular mechanisms essentially contribute to severe disease progression and mortality. High-resolution single-cell-transcriptomic analyses focused almost exclusively on immune cells and proposed treatment-relevant signatures (16). Therefore, we aimed at highlighting the interface between the immune and endothelial systems in order to define signatures of severe COVID-19 with the potential to develop novel therapeutic strategies. In our study, we analyzed the cellular and humoral immune response in ICU and CONV, including a variety of plasma proteins as they serve as key mediators of inflammation and endothelial integrity.

In peripheral blood of ICU, we identified major changes in the distribution of immune cell populations with a massive expansion of granulocytes along with a severe lymphopenia. Even though lymphopenia has been described as hallmark for severe COVID-19 it remains elusive whether it is caused by tissue infiltration or destruction of lymphocytes (17). As indicated by the T cell signature of CONV, SARS-CoV-2 infection causes a differentiation of naïve into effector CD8^+^ T cells, further supported by the significant increase of EM CD8^+^ T cells over time. Thus, the time-dependent differentiation of several immune cell populations could explain the high variability among proportions and numbers of immune cells in ICU. Several changes like the differentiation to memory T and NK cells persisted also in CONV arguing for long-term effects that may contribute to Long-COVID.

The immune signature of COVID-19 ICU displayed a highly activated phenotype, particularly of T and NK cells characterized, by a HLA-DR^+^CD38^+^ and CD127^-^ phenotype. Up to 80% of CD8^+^ T cells in ICU were HLA-DR^+^CD38^+^, demonstrating strong T cell activation in response to SARS-CoV-2. The appearance of HLA-DR^+^CD38^+^CD8^+^ T cells has also been described for other viruses like HIV and Influenza A and was associated with worse outcomes and impaired T cell function (18,19). For COVID-19 others have shown that conversely to their activated phenotype HLA-DR^+^CD38^+^CD8^+^ T cells also expressed exhaustion markers like PD-1 but were still functional by producing e.g. IFN-*γ*, granzyme B and perforin, suggesting that these cells are not generally functionally impaired by their PD-1 expression (20-23).

Regarding the humoral immunity to SARS-CoV-2, several studies demonstrated an extraordinary B cell response with absence of germline centers in deceased COVID-19 patients, appearance of extra-follicular B cell activation together with neutralizing antibodies, an extensive class switching to IgG and IgA accompanied by limited somatic hypermutation, as well as TGF-β instructed SARS-CoV-2 unspecific B cell responses (24-27). As others have previously shown (6), we observed a significant expansion of plasmablasts in severe COVID-19 patients accompanied by IgA and IgG SARS-CoV-2-specific antibodies. In our study, ICU not only produced high levels of S-and N-specific antibodies but the majority of these antibodies also blocked the binding of SARS-CoV-2 RBD to ACE2 *in vitro*, indicating the development of functional antibodies. Nevertheless, these patients developed severe COVID-19 symptoms despite the presence of blocking antibodies, suggesting that the humoral immune response alone may be insufficient to prevent critical disease progressions. As the development of SARS-CoV-2 specific antibodies in ICU together with the activation and differentiation of T and NK cells proves that these patients are immunocompetent, other mechanisms are likely to be involved in severe COVID-19 progression.

In our study, we observed that severely ill COVID-19 patients were characterized by a massive release of several plasma proteins indicating an endothelial damage and a previously reported COVID-19 associated cytokine storm (28). We identified seven core plasma proteins i.e. CXCL8-10, HGF, IL-6, IL-12(p40) and SCGF-*β* for severe COVID-19 that were associated with organ failure (SOFA-score), inflammation (CRP-level) and lung dysfunction (WHO-score). Furthermore, elevated levels of IGFBP-1, M-CSF, VCAM-1 and IL-16 correlated with high SOFA-and WHO-scores, indicating a connection of these soluble mediators to multiple organ and endothelial injuries. Overall, markedly elevated levels of these plasma proteins depict a highly inflammatory environment as well as endothelial activation and disruption and therefore are possible predictors for disease progression and outcome.

CXCL8-10 as well as IL-6 and IL-12(p40) are highly pro-inflammatory mediators and elevated in cytokine storm release syndrome and systemic diseases (28-30). IL-6 serves as a major orchestrator for systemic inflammation through induction of the hepatic acute phase response. Furthermore, high circulating levels of IL-6 as in a COVID-19-associated cytokine storm result in *trans* signaling allowing IL-6 together with sIL-6Rα to activate cells not expressing the membrane-bound IL-6R, like endothelial cells (28). As a result of the IL-6 mediated activation E-cadherin expression is reduced on endothelial cells leading to vascular hyperpermeability. Moreover, not only IL-6 but also HGF has been reported to directly contribute to endothelial disruption and vascular leakage (31). IGFBP-1 and its ligand IGF-1 are known to be elevated in severe lung injuries like idiopathic pulmonary fibrosis and ARDS (32,33). The fact that elevated IGFBP-1 levels are associated with hypoxia further suggests that IGFBP-1 may be a predictor for lung dysfunction and tissue damage (34,35). Since soluble VCAM-1 and M-CSF are additional indicators of endothelial activation and were identified as hallmark plasma proteins for severe COVID-19 (36,37), we propose that endothelial activation and disruption are the main drivers of severe COVID-19, which has not been demonstrated in parallel to immune dysregulation before.

The different plasma protein profiles from ICU suggest a crucial role of endothelial damage in severe COVID-19 and link the inflammatory response to endothelial barrier disruption. Focusing on ICU, unsupervised cluster analyses identified three subgroups based on their plasma protein profiles and implied a development from a primarily inflammatory phenotype in Sub1 to additional endothelial activation and disruption in Sub3 and an endothelial reconstitution and decline of inflammation in Sub2. A highly inflammatory phenotype with elevated plasma levels of IL-6, IL-12(p40), IFN-*γ* and CXCL10 characterized Sub1 patients without signs of endothelial damage. In contrast, remarkably high levels of HGF, Ang-2, OPN and VCAM-1 in Sub3 represented extended endothelial activation and disruption in addition to inflammation also mediated by IL-6, CXCL10, IL-18 and IL-12(p40). Furthermore, elevated secretion of key regulators of fibrinolysis like uPA and PAI-1 were unique features of Sub3 and illustrated a dysregulation of fibrinolysis and coagulation in addition to endothelial damage and inflammation. Since micro-thrombotic patterns are well described for severe COVID-19 (38) and represent a serious complication, these results support the assumption of Sub3 as high-risk patients with systemic inflammation, tissue damage and impaired coagulation and therefore poor survival prospects.

Sub2 was characterized by lower levels of pro-inflammatory cytokines and endothelial factors like IL-6, IL-12(p40), uPA, HGF and IGFBP-1, together with decreased SOFA-and WHO-scores suggesting a decline in inflammation and endothelial damage. Furthermore, Sub2 exhibited the highest levels of the anti-inflammatory cytokines IL-13 and IL-1 which correlated with decreased SOFA-and WHO-scores as well as CRP levels. While IL-13 was reported to be required for recovery after acute lung injury by regulating local levels of IL-6, CCL2/MCP-1 and G-CSF, IL-1*β* is usually known as a pro-inflammatory cytokine related to ARDS (39,40). However, other studies propose that IL-1*β* is involved in tissue healing and has beneficial effects on endothelial repair after ARDS (41-43). Additionally, GM-CSF and VEGF are further factors elevated in Sub2 and significantly increased over time together with IL-1*β* and IL-13. Correspondingly, GM-CSF and VEGF also have been described to essentially contribute to tissue repair and wound healing (44-46). These observations indicate that elevated levels of IL-1*β*, IL-13, GM-CSF and VEGF after 20 days of symptom onset are predictors for tissue repair and improved health condition of severe COVID-19 patients. The observation that Long-COVID syndrome is characterized by an impairment of several organ systems including pulmonary, cardiovascular and neurophysiological symptoms (13) further suggests a continuous contribution of the endothelial-epithelial barrier to Long-COVID. Therefore, knowing the mechanisms involved in establishment and recovery of COVID-19, focusing especially on the endothelium, may be essential for more precise diagnostics for Long-COVID and even treatment strategies.

In conclusion, we propose that in severe cases SARS-CoV-2 infection leads to a massive local inflammation developing into a dysregulated immune activation with cytokine and chemokine release. The combination of this inflammation with a systemic endothelial activation can proceed towards capillary leakage and multi-organ failure. The limitations of our study include the single-center setting with a rather small and heterogeneous patient cohort. Furthermore, samples were obtained at variable time points during disease. Further studies are needed to define therapeutic consequence of our observations. To the best of our knowledge, the present study shows for the first time that endothelial damage is another major driver of COVID-19 severity together with substantial immune dysregulation. Thus, severe and life-threatening conditions of COVID-19 patients are not only characterized by a highly activated immune phenotype and pro-inflammatory cascades but also by substantial endothelial injuries, which may explain multi-organ involvement in severe COVID-19.

## Materials and Methods

### Study design

In total 25 patients from intensive care unit (ICU) and 17 convalescent patients were recruited to this study between April and September 2020 at Hannover Medical School (MHH). Convalescent cohort included former ICU patients. ICU and CONV had a confirmed SARS-CoV-2 infection via viral PCR. For some patients, samples were taken at multiple time points leading to a sample size of 58 for ICU patients and 28 for convalescent patients. Five patients from ICU died and were visualized with the last sample obtained before decease. Clinical severity of COVID-19 was assessed on different parameters. World Health Organizations (WHO) eight-point scale for COVID-19 trail endpoints was used to classify severity of COVID-19 (https://www.who.int/publications/i/item/covid-19-therapeutic-trial-synopsis). C-reactive protein (CRP) level was used as inflammation marker, whereas sepsis-related organ failure assessment (SOFA) score determined the extent of systemic organ failure. PF ratio represented the individual pulmonary function. Clinical and demographical characteristics of study participants are summarized in Supplementary Table 1. Of the 25 ICU patients, 12 received Remdesivir and 6 received Tocilizumab treatment. 29 unexposed individuals (UE) were enrolled as control group, some also at several time points leading to a sample size of 36 for UE.

### Quantification of cells from EDTA blood via TruCount™ analysis

Absolute cell numbers were calculated from whole blood using BD Trucount™ Tubes (BD Biosciences), following manufacturer’s instructions.

### Flow cytometry

For flow cytometry analyses, whole blood EDTA samples (100 µl) were incubated with antibodies for surface staining in FACS Buffer (0,1% NaN3, 2,5% FCS in PBS) at 4°C for 30 min and followed by 15 min erythrocyte lysis using 1x BD Lysing Solution. Cells were washed with PBS prior to acquisition. All antibodies used for flow cytometry analyses are listed in Supplementary Table 2. Cells were acquired and analyzed on a LSRII flow cytometer (BD Biosciences, USA) using FACS Diva software (v8.0).

### Multiplex assays

Luminex-based multiplex assays were used to quantify plasma proteins and SARS-CoV-2 S-and N-specific antibodies.

Plasma proteins were measured using the Bio-Plex Pro^TM^ Human Assays (Bio-Rad, Hercules, USA): cytokine screening panel plus ICAM-1 and VCAM-1 (12007283, 171B6009M, 171B6022M), Cancer Biomarker Panel 1 and Panel 2 (171AC500M, 171AC600M) and Th17 cytokine Panel 15-Plex (171AA001M) following manufacturer’s instructions with thawed plasma and which was diluted twofold with assay buffer.

SARS-CoV-2 S-and N-specific antibodies were detected using the SARS-CoV-2 Antigen Panel 1 IgG, IgM, IgA assay (Millipore HC19SERM1-85K-04, HC19SERA1-85K-04, HC19SERG1-85K-04) following manufacturer’s instructions. The assays were performed with thawed plasma, which was diluted 1:200 for ICU and CONV and 1:100 for UE with assay buffer.

The semi-quantitative readout is given as median fluorescence intensity (MFI) of > 50 beads for each antigen and sample, acquired by the Bio-Plex 200 machine and the Bio-Plex Manager^TM^ Version 6.0 software (Bio-Rad Hercules, USA).

### Competitive ELISA

Competitive ELISA was performed to detect blocking antibodies against SARS-CoV-2 RBD. Plates were coated over night at 4°C with 50 µl/well of recombinant RBD (400 ng/ml). After 3 times washing, plates were blocked for 1-2 h at room temperature (RT) with PBS+2%FCS. 0,25 µg/ml biotinylated human ACE2 (Acro Biosystems) were added followed by 50 µl patient plasma (1:50 dilution). Plates were incubated at RT for 1-2 h and washed 3 times afterwards. HRP-bound streptavidin (Merck-Millipore) was added and incubated at RT for 1-2 h, then washed 3-4 times. TMB substrate (BD Biosciences) was added and reaction was stopped with 50 µl H2SO4 (0,5 M). Optical densities were measured at 450 nm. Efficient blocking was expressed as the percent neutralization at a 1:50 plasma dilution relative to a UE control serum.

### Statistical analyses

Statistical analyses of the data were performed with GraphPad Prism v7.0/v9.0 software (GraphPad Software). D’Agostino-Pearson omnibus normality test was calculated to assess data distribution. Parametric tests were performed where data were normally distributed, otherwise non-parametric tests were used. The statistical test used in each analysis is indicated in the figure legends. Correlation analyses were performed using Spearman rank-order correlation. Results were considered significant if p<0.05.

Qlucore Omics Explorer (version 3.6, Qlucore) was used to generate principal component analysis (PCA) plots, heatmaps, tSNE plots and volcano plots. For each analysis, q-value or variance used as a cut-off are indicated in the figure legends.

For the cytokine correlation matrix Spearman rank-order correlation coefficient was visualized using the corrplot R package.

### Study approval

The study was approved by the Hannover Medical School Ethics Committee. All patients or participants provided written informed consent before participation in the study (9001 BO K, 968-2011).

## Data Availability

All data associated with this study are available in the main text or the supplementary materials.

## Author contribution

LR performed flow cytometry experiments, analyzed data, prepared figures and wrote the manuscript. IP collected clinical data, provided patient samples and reviewed the manuscript. JFK performed flow cytometry experiments and supported writing the manuscript. KB and JK performed multiplex experiments. SC performed flow cytometry experiments. ER performed competitive ELISA experiments and HMJ helped interpreting data and edited the manuscript. TFS reviewed and edited the manuscript. MC, TW and MMH provided patient samples. AS, LB, JS and SD collected samples. CSF supervised and designed the study and wrote the manuscript.

## Acknowledgments

This project was supported by the German Research Foundation DFG FA-483/1-1, the German Center for Infection Research DZIF TTU-IICH 07_913 and the Lower Saxony Ministry of Research and Culture (ImProVIT). HMJ is supported by a grant (01KI2043) from the Bundesmininisterium für Bildung und Forschung (BMBF), funds from the Bavarian State ministry for Science and the BMBF-funded COVIM project (NaFoUniMedCovid19).

