## Supplemental Figures for "Endothelial dysfunction determines severe COVID-19 in combination with dysregulated lymphocyte responses and cytokine networks"

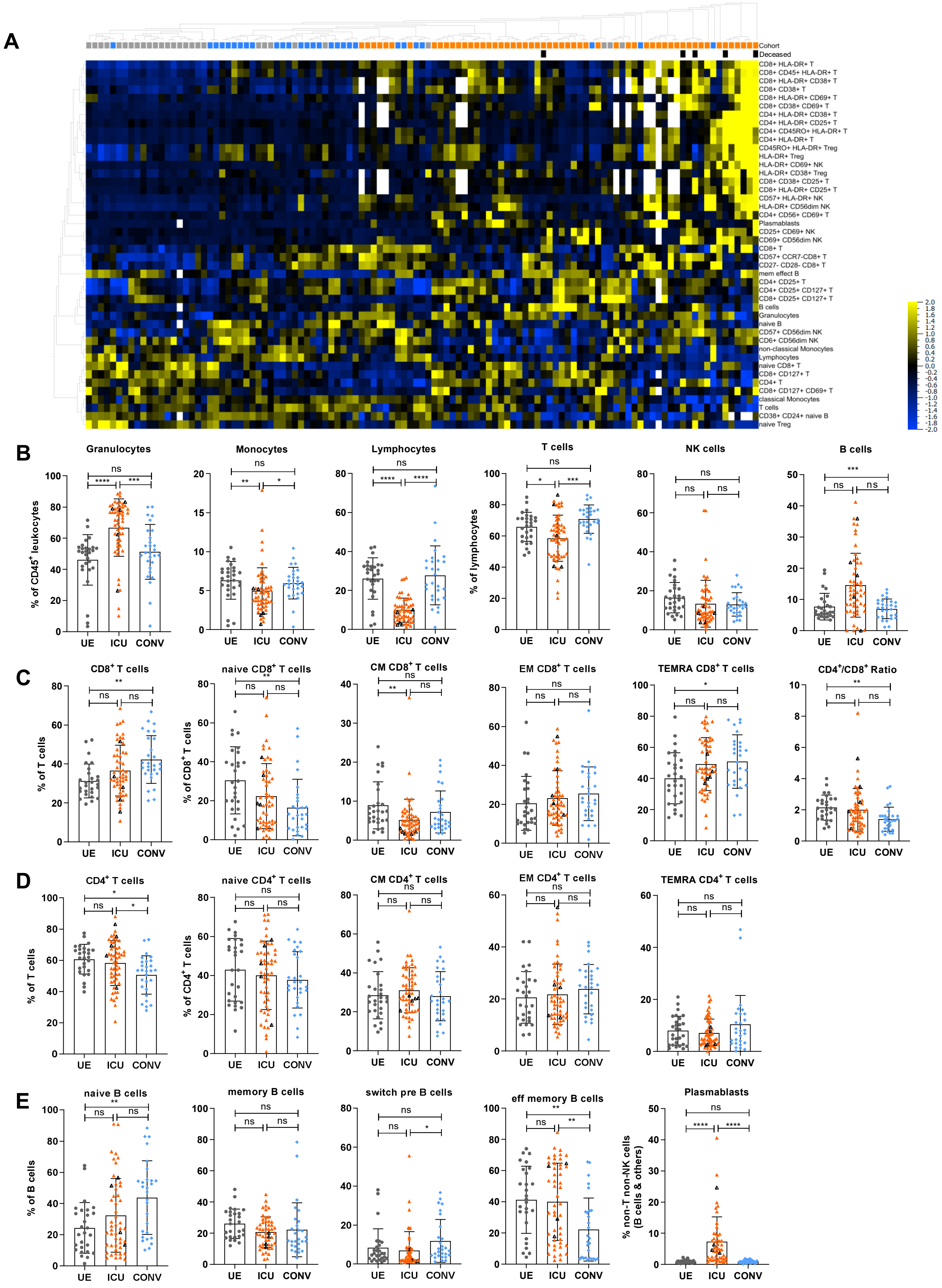
A
Cohort
Deceased
CD8+ HLA-DR+ T
CD8+ CD45+ HLA-DR+ T
CD8+ HLA-DR+ CD38+ T
CD8+ CD38+ T
CD8+ HLA-DR+ CD69+ T
CD8+ CD38+ CD69+ T
CD4+ HLA-DR+ CD38+ T
CD4+ HLA-DR+ CD25+ T
CD4+ CD45RO+ HLA-DR+ T
CD4+ HLA-DR+ T
CD45RO+ HLA-DR+ Treg
HLA-DR+ Treg
HLA-DR+ CD69+ NK
HLA-DR+ CD38+ Treg
CD8+ CD38+ CD25+ T
CD8+ HLA-DR+ CD25+ T
CD57+ HLA-DR+ NK
HLA-DR+ CD56dim NK
CD4+ CD56+ CD69+ T
Plasmablasts
CD25+ CD69+ NK
CD69+ CD56dim NK
CD8+ T
CD57+ CCR7-CD8+ T
CD27- CD28- CD8+ T
mem effect B
CD4+ CD25+ T
CD4+ CD25+ CD127+ T
CD8+ CD25+ CD127+ T
B cells
Granulocytes
naive B
CD57+ CD56dim NK
CD6+ CD56dim NK
non-classical Monocytes
Lymphocytes
naive CD8+ T
CD8+ CD127+ T
CD4+ T
CD8+ CD127+ CD69+ T
classical Monocytes
T cells
CD38+ CD24+ naive B
naive Treg
B
Granulocytes
Monocytes
Lymphocytes
T cells
NK cells
B cells
% of CD45+ leukocytes
% of lymphocytes
C
CD8+ T cells
naive CD8+ T cells
CM CD8+ T cells
EM CD8+ T cells
TEMRA CD8+ T cells
CD4+/CD8+ Ratio
% of T cells
% of CD8+ T cells
D
CD4+ T cells
naive CD4+ T cells
CM CD4+ T cells
EM CD4+ T cells
TEMRA CD4+ T cells
% of CD4+ T cells
E
naive B cells
memory B cells
switch pre B cells
eff memory B cells
Plasmablasts
% of B cells
% non-T non-NK cells (B cells & others)
UE
ICU
CONV

**Fig. S1: Altered frequencies of certain immune cells in severe COVID-19 patients.**

Immune cell distribution reflected by frequencies in patient blood was analyzed using flow cytometry. **(A)** Heatmap analysis. Multigroup comparison with a p-value cut off of 0.041 was used to identify significant differences among UE, ICU and CONV. **(B-E)** Frequencies of different immune cells in patient blood. T cells: naïve (CCR7+CD45RO-), central memory (CM, CCR7+CD45RO+), effector memory (EM, CCR7-CD45RO+) and TEMRA (CCR7-CD45RO-); B cells: naïve (IgD+CD27-) memory (mem, CD27+IgD-), switch precursor (switch pre, CD27+IgD+), effector memory (eff mem, IgD-CD27-) and plasmablasts (CD19+CD20-CD27+CD38+)

Black triangles represent last samples from deceased patients.

UE: Unexposed donors, ICU: Intensive care unit patients, CONV: convalescent patients, CM: central memory, EM: effector memory, mem eff: memory effector

Statistical analysis: ANOVA test with Turkey multiple comparison test or Kruskal-Wallis with test with Dunn's multiple comparison test were performed. *$p < 0.05$, **$p<0.01$, ***$p<0.001$, ****$p<0.0001$.

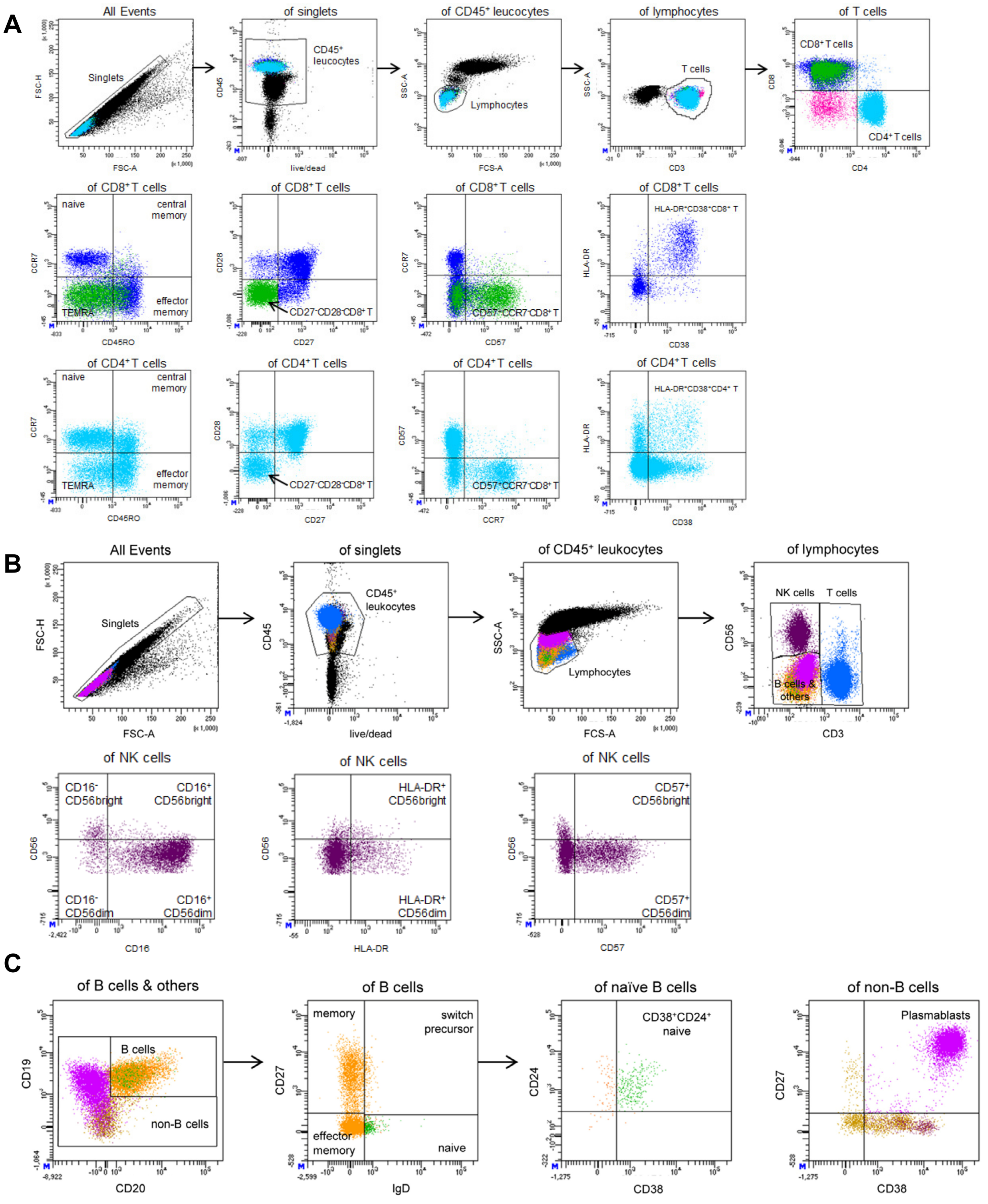


**Fig. S2: Flow cytometry gating strategy.**

Representative flow cytometry plots visualizing the gating strategy for **(A)** T cell subsets, **(B)** NK cell subsets and **(C)** B cell subsets.

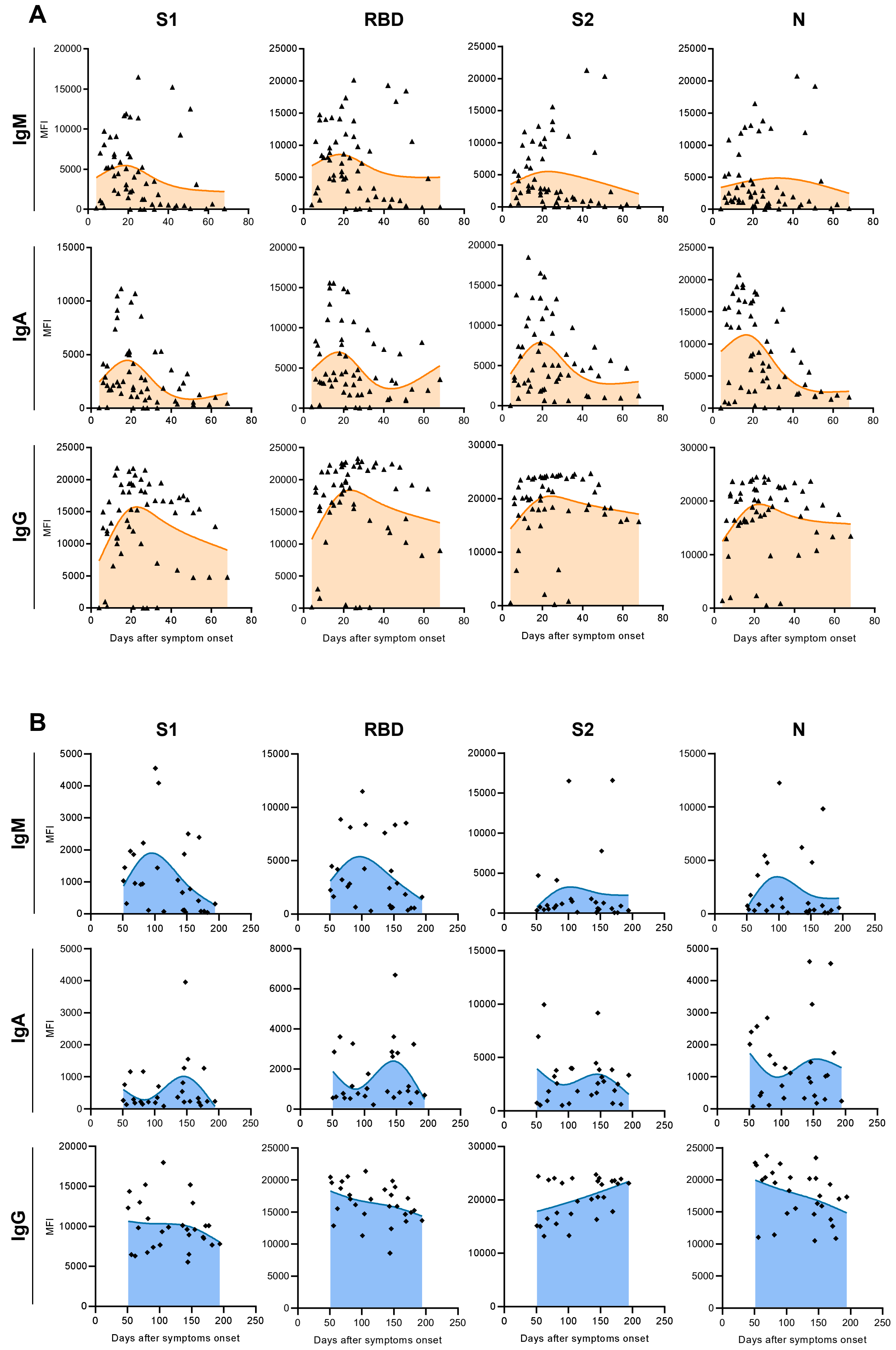

A
S1
RBD
S2
N
IgM
IgA
IgG
MFI
Days after symptom onset
B
Days after symptoms onset

**Fig. S3: Changes in SARS-CoV-2 specific antibody levels over time in ICU and CONV.**

Luminex-based multiplex assay was used to detect SARS-CoV-2 specific IgM, IgA and IgG antibodies against S1-, S2-, N- or RBD-antigen of SARS-CoV-2 in patient sera. Antibody levels were plotted against days after symptom onset for **(A)** ICU and **(B)** CONV.

Curves were generated with LOWESS regression.

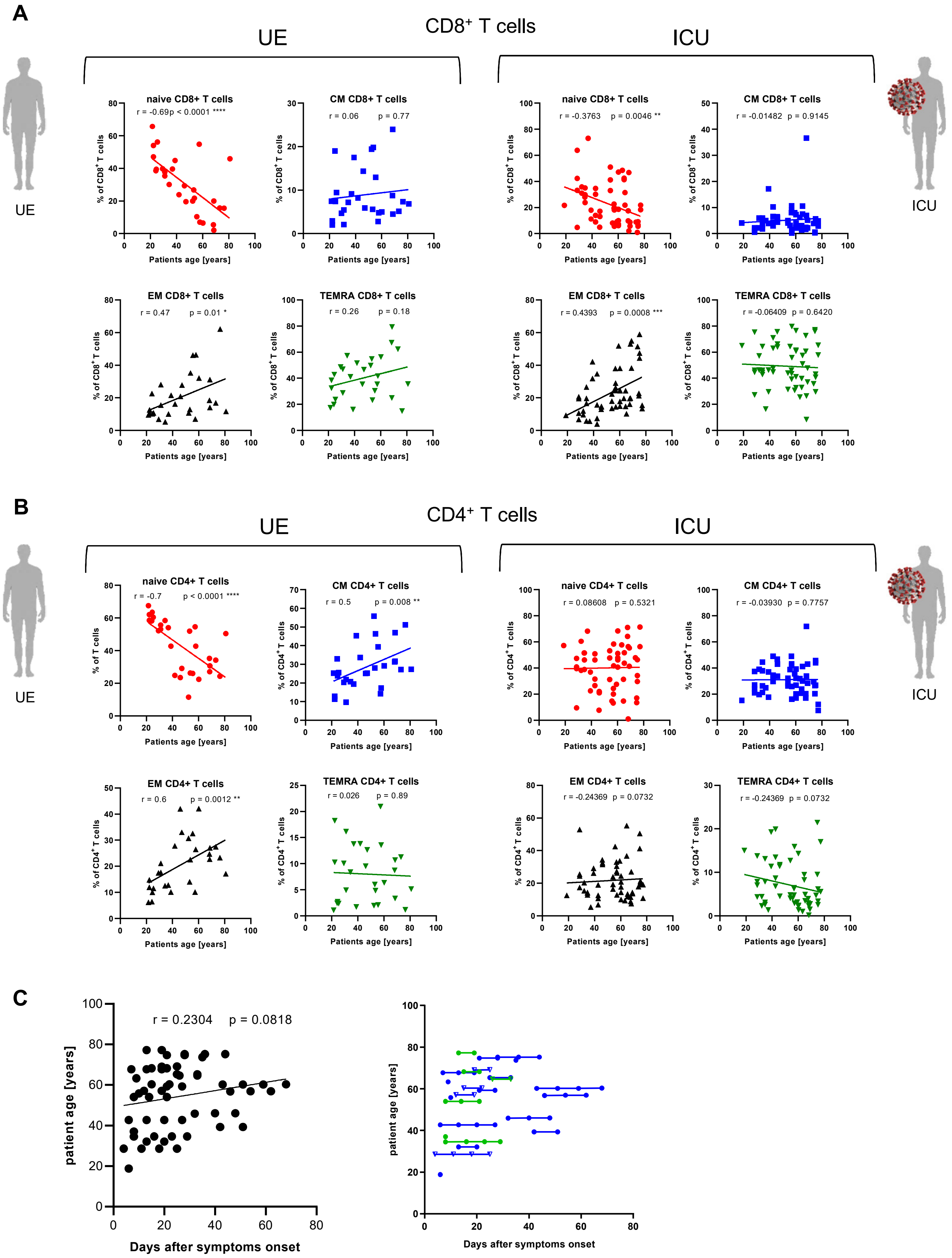
A
CD8+ T cells
UE
ICU
naive CD8+ T cells
r = -0.69p < 0.0001 ****
CM CD8+ T cells
r = 0.06 p = 0.77
naive CD8+ T cells
r = -0.3763 p = 0.0046 **
CM CD8+ T cells
r = -0.01482 p = 0.9145
EM CD8+ T cells
r = 0.47 p = 0.01 *
TEMRA CD8+ T cells
r = 0.26 p = 0.18
EM CD8+ T cells
r = 0.4393 p = 0.0008 ***
TEMRA CD8+ T cells
r = -0.06409 p = 0.6420
% of CD8+ T cells
Patients age [years]
B
CD4+ T cells
UE
ICU
naive CD4+ T cells
r = -0.7 p < 0.0001 ****
CM CD4+ T cells
r = 0.5 p = 0.008 **
naive CD4+ T cells
r = 0.08608 p = 0.5321
CM CD4+ T cells
r = -0.03930 p = 0.7757
EM CD4+ T cells
r = 0.6 p = 0.0012 **
TEMRA CD4+ T cells
r = 0.026 p = 0.89
EM CD4+ T cells
r = -0.24369 p = 0.0732
TEMRA CD4+ T cells
r = -0.24369 p = 0.0732
% of T cells
% of CD4+ T cells
Patients age [years]
C
r = 0.2304 p = 0.0818
patient age [years]
Days after symptoms onset

**Fig. S4: Age-related changes in T cell subsets in ICU patients.**

**(A, B)** Frequencies of $CD8^+$ and $CD4^+$T cell subsets in ICU patient blood, including naïve (CCR7+CD45RO-), central memory (CM, CCR7+CD45RO+), effector memory (EM, CCR7-CD45RO+) and TEMRA (CCR7-CD45RO-), were analyzed using flow cytometry. Frequencies were correlated to patient age. **(C)** Correlation between patient age and days after symptom onset (left). ICU patient age against days after symptom onset. Repeated samples from the same individuum are linked. Green dots represent samples from female patients. Blue dots represent samples from male patients. Triangles represent samples from deceased patients.

Statistical analysis: Spearman-Correlation. *$p < 0.05$, **$p<0.01$, ***$p<0.001$, ****$p<0.0001$.

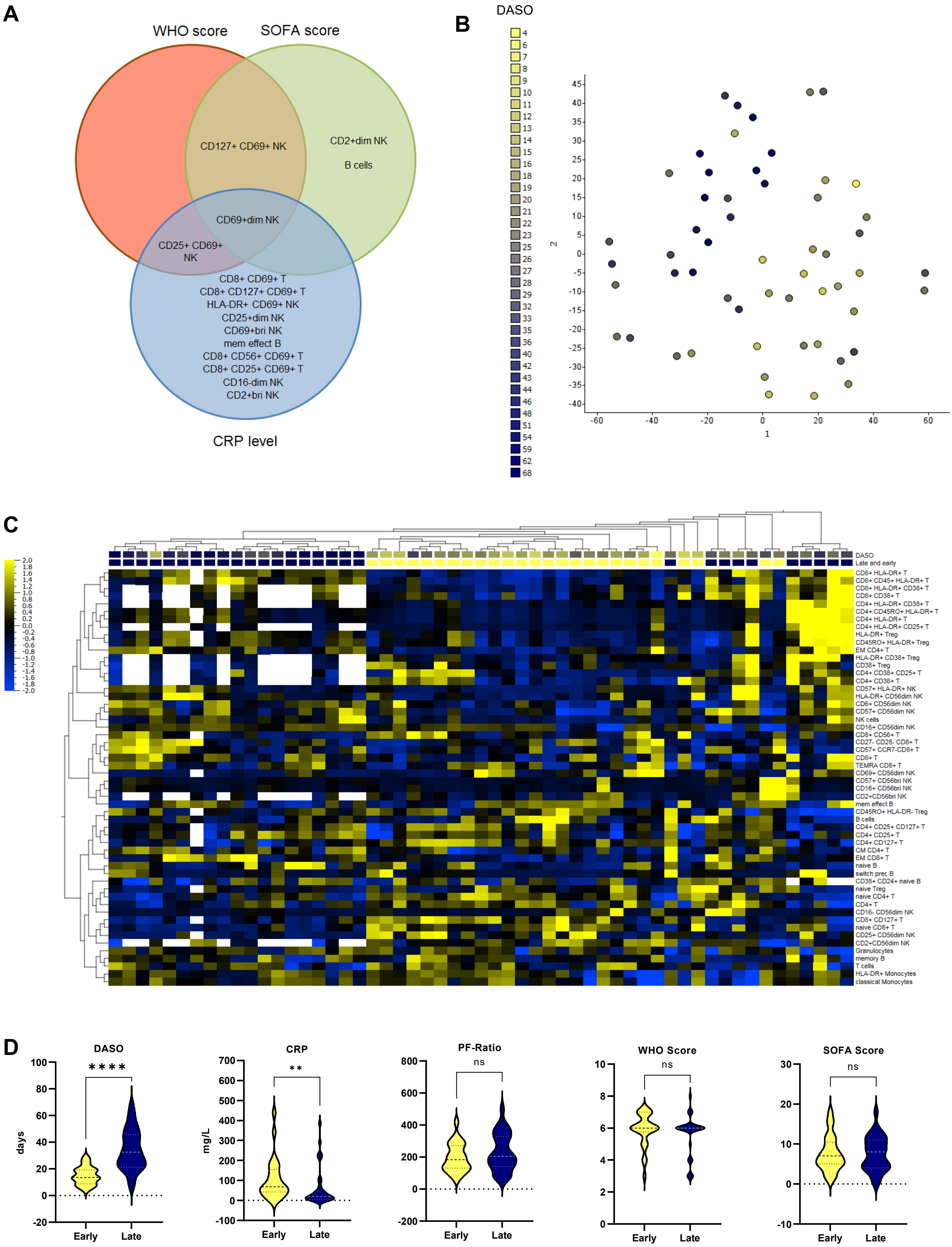
A
WHO score
SOFA score
CD127+ CD69+ NK
CD2+dim NK
B cells
CD69+dim NK
CD25+ CD69+ NK
CD8+ CD69+ T
CD8+ CD127+ CD69+ T
HLA-DR+ CD69+ NK
CD25+dim NK
CD69+bri NK
mem effect B
CD8+ CD56+ CD69+ T
CD8+ CD25+ CD69+ T
CD16-dim NK
CD2+bri NK
CRP level
B
DASO
C
DASO
Late and early
CD8+ HLA-DR+ T
CD8+ CD45+ HLA-DR+ T
CD8+ HLA-DR+ CD38+ T
CD8+ CD38+ T
CD4+ HLA-DR+ CD38+ T
CD4+ CD45RO+ HLA-DR+ T
CD4+ HLA-DR+ T
CD4+ HLA-DR+ CD25+ T
HLA-DR+ Treg
CD45RO+ HLA-DR+ Treg
EM CD4+ T
HLA-DR+ CD38+ Treg
CD38+ Treg
CD4+ CD38+ CD25+ T
CD4+ CD38+ T
CD57+ HLA-DR+ NK
HLA-DR+ CD56dim NK
CD6+ CD56dim NK
CD57+ CD56dim NK
NK cells
CD16+ CD56dim NK
CD8+ CD56+ T
CD27- CD28- CD8+ T
CD57+ CCR7-CD8+ T
CD8+ T
TEMRA CD8+ T
CD69+ CD56dim NK
CD57+ CD56bri NK
CD16+ CD56bri NK
CD2+CD56bri NK
mem effect B
CD45RO+ HLA-DR- Treg
B cells
CD4+ CD25+ CD127+ T
CD4+ CD25+ T
CD4+ CD127+ T
CM CD4+ T
EM CD8+ T
naive B
switch prec B
CD38+ CD24+ naive B
naive Treg
naive CD4+ T
CD4+ T
CD16- CD56dim NK
CD8+ CD127+ T
naive CD8+ T
CD25+ CD56dim NK
CD2+CD56dim NK
Granulocytes
memory B
T cells
HLA-DR+ Monocytes
classical Monocytes
D
DASO
****
days
Early
Late
CRP
**
mg/L
Early
Late
PF-Ratio
ns
Early
Late
WHO Score
ns
Early
Late
SOFA Score
ns
Early
Late

**Fig. S5: Correlation between the immune cell composition of COVID-19 ICU patients and disease progression**

**(A)** Venn-diagram displaying significantly positive correlations between immune cell subsets and CRP levels, SOFA- and WHO-score **(B)** tSNE analysis of flow cytometry data from ICUs. Variance-value cut off of 0.305 was used to identify significant differences within the ICU cohort. Patients samples were colored based on their days after symptom onset. **(C)** Heatmap of flow cytometry data including 98 immune cell populations from ICUs. Variance-value cut off of 0.035 was used to identify significant differences within the ICU cohort. Samples and immune cell subsets were ordered according to hierarchical clustering. Patients were classified into two groups. Blue: Late subgroup, yellow: early subgroup. Blue to yellow scale indicates the prevalence of each subset. Missing values are displayed in white. **(D)** Clinical parameters from early and late ICU patients.

DASO: Days after symptom onset

Statistical analysis: Unpaired t-test or Mann-Whitney test. *$p < 0.05$, **$p<0.01$, ***$p<0.001$, ****$p<0.0001$.

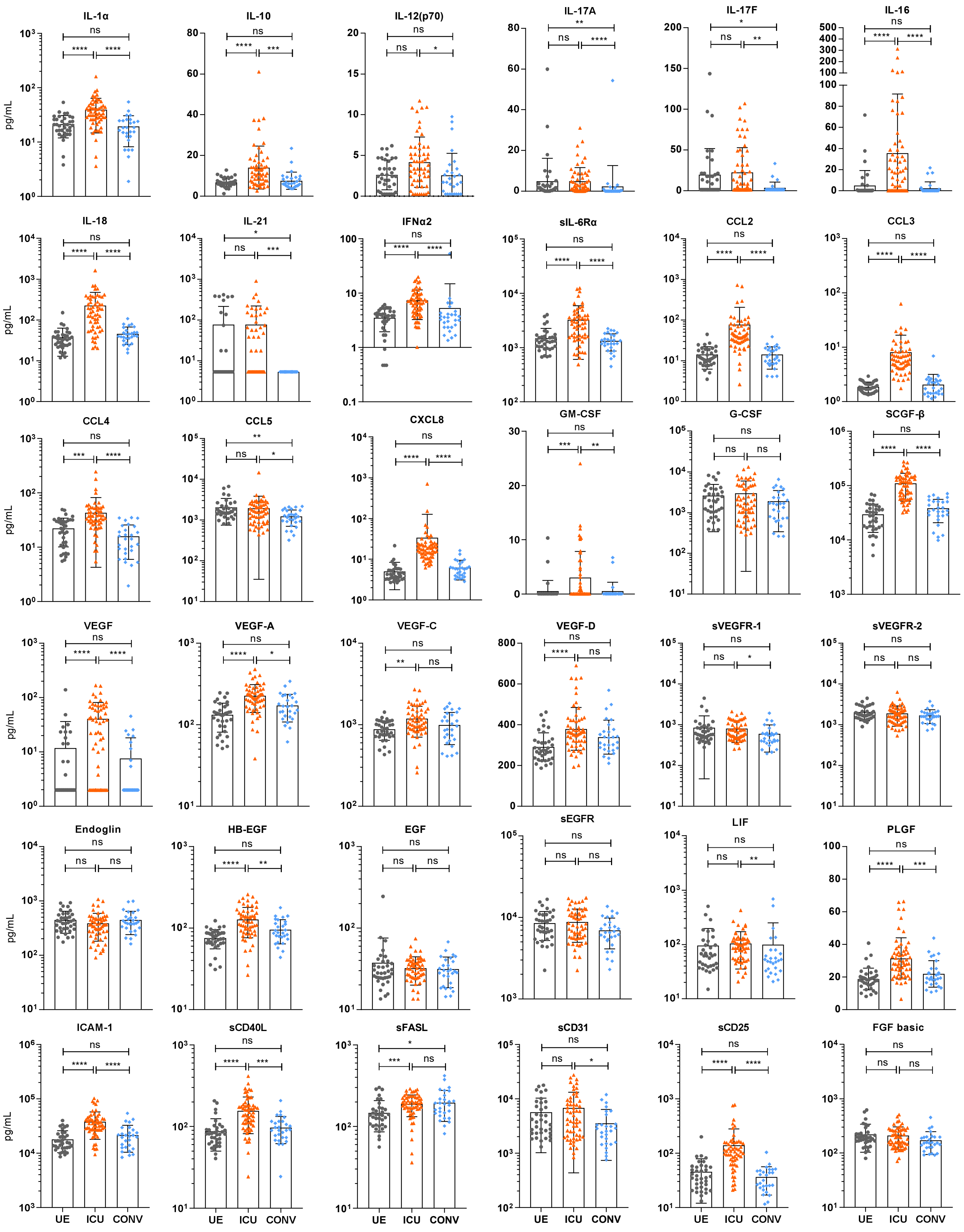


**Fig. S6: Levels of cytokines, chemokines and endothelial factors in plasma of UE, ICU and CONV.**

Luminex-based multiplex assay was used to analyze levels of cytokines and chemokines in plasma from UE, ICU and CONV patients.

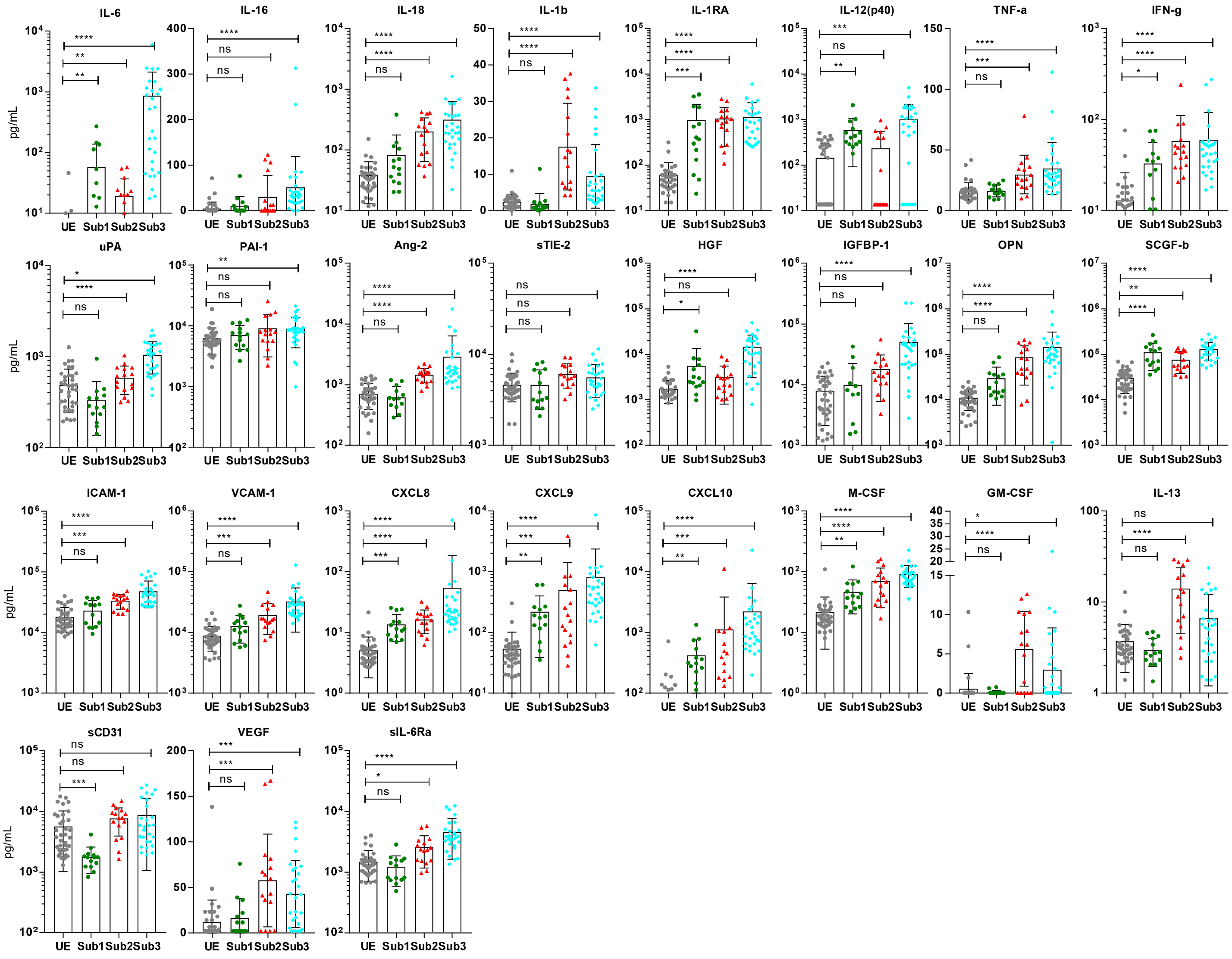


**Fig. S7: Differences in plasma proteins levels among ICU subgroups compared to UE.**

Luminex-based multiplex assay was used to analyze plasma protein levels in UE and ICU patients. ICU patients were classified into three subgroups (Sub1, Sub2, Sub3).

Statistical analysis: Kruskal-Wallis test with test with Dunn's multiple comparison test were performed. *$p < 0.05$, **$p<0.01$, ***$p<0.001$, ****$p<0.0001$.

**Table S1: Patients demographics and clinical characteristics.**
Characteristics of intensive care unit patients (ICU), convalescent patients (CONV) and unexposed donors (UE). N=number of patients, n=number of samples.

| **Characteristics** | **ICU cohort** N=25 | **CONV cohort** N=17 | **UE cohort** N=29 |
|---|---|---|---|
| **Samples number n** | 58 | 28 | 36 |
| **Age** in years (Min - Max) | 54,2 (18,8 - 77,2) | 53,3 (18,9 – 75,5) | 45,3 (21,5 – 80,9) |
| **Gender** | | | |
| male | 44 (76%) | 22 (79%) | 15 (42%) |
| female | 14 (24%) | 6 (21%) | 21 (58%) |
| **COVID risk factors/Co-morbidities** | | | |
| Chronic lung disease | 3 (2) | 7 (4) | 0 |
| Chronic cardiovascular disease | 8 (4) | 6 (4) | 1 (1) |
| Chronic renal failure | 4 (1) | 1 (1) | 0 |
| Chronic liver disease | 0 | 2 (1) | 0 |
| Cancer | 6 (3) | 1 (1) | 0 |
| Lymphoma | 2 (1) | 0 | 0 |
| Pancreas | 3 (1) | 1 (1) | 0 |
| NSCLC | 1 (1) | 0 | 0 |
| Diabetes | 14 (8) | 5 (4) | 5 (3) |
| Adiposis | 33 (15) | 8 (5) | 5 (3) |
| Hypertension | 28 (14) | 11 (7) | 7 (5) |
| Pregnancy | 5 (2) | 2 (2) | 0 |
| Immunosuppression | 8 (3) | 5 (3) | 0 |
| Solid Organ Transplant | 4 (1) | 1 (1) | 0 |
| HIV | 0 | 2 (1) | 0 |
| Other | 4 (2) | 2 (1) | 0 |
| **Clinical parameter** | | | |
| DASO (Mean, Min, Max) | 25,5 (4 - 68) | 120 (51 – 194) | / |
| CRP mg/L (Mean, Min, Max) | 83,79 (2,2 - 439) | / | / |
| PF ratio (Mean, Min, Max) | 209,3 (62 - 500) | / | / |
| SOFA score (Mean, Min, Max) | 7,5 (0 - 18) | / | / |
| WHO score (Mean, Min, Max) | 6 (3 - 8) | / | / |
| **MIS-C with COVID-19** | 1 (1) | 0 | / |
| **Deceased** | 5 (20%) | / | / |

NSCLC: Non-small-cell lung carcinoma, DASO: Days after symptom onset, MIS-C: Multisystem Inflammatory Syndrome in Children

**Table S2: Flow cytometry antibodies.**
Antibodies used for flow cytometry analyses.

| Antigen | Flurochrome | Manufacturer |
|---|---|---|
| CD3 | V500 | BD Bioscience |
| CD3 | APC-H7 | BD Bioscience |
| CD3 | PerCP | BD Bioscience |
| CD4 | PerCP | BD Bioscience |
| CD6 | FITC | BD Bioscience |
| CD8 | APC-H7 | BD Bioscience |
| CD14 | PE-Cy7 | BD Bioscience |
| CD16 | APC | BD Bioscience |
| CD19 | PerCP | BD Bioscience |
| CD20 | APC-H7 | BD Bioscience |
| CD24 | FITC | BD Bioscience |
| CD25 | BV421 | BD Bioscience |
| CD27 | FITC | BD Bioscience |
| CD27 | BV421 | BD Bioscience |
| CD28 | APC | BD Bioscience |
| CD38 | APC | BD Bioscience |
| CD45 | AF700 | Biolegend |
| CD45 | APC-H7 | BD Bioscience |
| CD45R0 | PE-Cy7 | BD Bioscience |
| CD56 | PE | BD Bioscience |
| CD57 | BV421 | BD Bioscience |
| CD69 | FITC | BD Bioscience |
| CD127 | AF647 | BD Bioscience |
| CD197 (CCR7) | PE | BD Bioscience |
| HLA-DR | V500 | BD Bioscience |
| IgD | PE-Cy7 | BD Bioscience |
